# Lenalidomide enhances CD19 CAR-T cell fitness and target-cell engagement in relapsed/refractory CLL

**DOI:** 10.64898/2026.06.23.26356089

**Authors:** Mikalai Katsin, Valeria M. Stepanova, Dmitri Dormeshkin, Alexandr Migas, Dzmitry Lutskovich, Alexander Meleshko, Yuliya Serada, Yauheniya Khalankova, Tatsiana Shman, Hanna Klych, Kate Lutskovich, Elizaveta R. Naberezhnaya, Andrei S. Logvinov, Dmitrii Pershin, Katerina Malahova, Victoryia Hrytsyva, Arina Trigorlova, Nina Velko, Hanna Kasyanenka, Michael A. Maschan, Alexander G. Gabibov, Veranika Bakhir, Anzhela Tomchyna, Anzhalika Solntcava, Alexey V. Stepanov

**Author notes:** These authors contributed equally to this work (Mikalai Katsin, Valeria M. Stepanova).

## Abstract

**Background:** CD19-directed CAR-T cell therapy can induce durable remissions in chronic lymphocytic leukemia (CLL), but response rates are lower than in other B-cell malignancies, in part because CLL is characterized by T-cell dysfunction, defective immune synapse formation, and impaired target-cell co-stimulation. Lenalidomide is an immunomodulatory drug with the potential to act on both sides of the CAR-T/CLL interface by improving T-cell fitness and modifying malignant B-cell susceptibility to immune engagement.

**Methods:** We are conducting an open-label, non-randomized phase I/II clinical trial VTB-CLL002 (ClinicalTrials.gov identifier: NCT06762431) evaluating the safety and efficacy of CD19 CAR-T cell therapy combined with concomitant lenalidomide in patients with relapsed or refractory CLL and small lymphocytic lymphoma followed by lenalidomide maintenance. The primary endpoint was safety. The secondary endpoint included overall response rate (ORR), complete response (CR), progression-free survival (PFS) and overall survival (OS).

**Results:** Twelve patients were treated. The median age was 60 years and the median number of prior lines of therapy was 2. All patients were BTK inhibitor-naive, and all had measurable disease at the time of infusion. CAR-T manufacturing was successful in all patients. All treated patients achieved complete remission, with a median time to response of 1 month. CAR T-cells expansion was observed in all patients, with a median peak expansion of 137 cells/µL and a median time to peak expansion of 14 days. CAR T-cells remained detectable at the last follow-up in all patients, with persistence documented up to 24 months. At dose levels 2–3, eight of nine patients had ongoing MRD-negative responses at the time of analysis.

Toxicity was clinically meaningful. Cytokine release syndrome (CRS) occurred in all patients, with severe CRS observed in 2 of 12 patients. ICANS occurred in 5 of 12 patients, including severe ICANS in 4 of 12 patients. One patient developed late grade 4 ICANS temporally associated with lenalidomide reintroduction and secondary CAR-T expansion. Early and late immune effector cell-associated hematotoxicity were common.

In mechanistic studies, lenalidomide enhanced CAR-T proliferation and cytotoxicity, shifted CAR-T cells toward effector-associated phenotypes, reduced selected exhaustion markers during repeated antigen challenge, and increased IL-2 and IFN-γ secretion. Lenalidomide also increased CAR-T/CLL conjugate formation and upregulated CD54/ICAM-1 on CLL target cells without broad induction of CD80, CD86, or CD40, consistent with improved adhesive target-cell engagement rather than classical co-stimulation. Transcriptomic profiling supported enhanced Th1/cytotoxic and T-cell activation-associated programs with lower T reg-associated genes in lenalidomide-treated CAR-T cells.

**Conclusions:** Lenalidomide-augmented CD19 CAR-T therapy demonstrated strong early clinical activity in relapsed/refractory CLL, characterized by deep responses, durable CAR-T persistence, and substantial incidence of immune effector-associated toxicities. These findings support further evaluation of lenalidomide as a rational CAR-T partner in CLL and suggest that its activity may involve both improved CAR-T fitness and enhanced target-cell engagement. Future studies should optimize lenalidomide timing and dosing to preserve response depth while reducing delayed immune-effector toxicity.

**Key points:** Key point 1

Lenalidomide may improve CD19 CAR-T activity in CLL by acting on both CAR-T cells and CLL cancer cells, enhancing immune synapse formation and effector function.

Key point 2

In 12 R/R CLL patients, combination therapy with lenalidomide and CD19 CAR-T cells demonstrated a high complete remission rate, frequent MRD negativity, and durable persistence, despite a high but manageable incidence of ICANS.

## Introduction

Chronic lymphocytic leukemia (CLL) and small lymphocytic leukemia (SLL) are mature B-cell neoplasms accounting for high incidence in Western countries. Over the past decade, Bruton tyrosine kinase inhibitors (BTKi) and BCL2 inhibitors (BCL2i) have significantly improved survival outcomes and reshaped the treatment landscape for CLL (1). However, the median overall survival for patients with double-refractory disease remains limited, reported to be only approximately 3.6 months (2). This unmet medical need continues to drive the development of new and innovative treatment approaches for CLL.

CD19-directed chimeric antigen receptor T-cell therapy has emerged as a therapeutic option for relapsed/refractory CLL/SLL. Lisocabtagene maraleucel, an autologous CD19 CAR-T product incorporating a 4-1BB costimulatory domain and a defined CD4:CD8 composition, has demonstrated clinically meaningful activity in patients previously treated with BTKi and BCL2i. However, responses in CLL remain less frequent than those observed in other B-cell malignancies (3), with reported overall and complete response rates of approximately 44% and 20%, respectively (4,5) The curative potential of CD19 CAR-T cells in CLL has been documented, underscoring the critical need for strategies that improve response depth and durability (6). The limited efficacy of CAR-T therapy in CLL is likely multifactorial. CLL is associated with intrinsic T-cell dysfunction, impaired immune activation, altered cytokine signaling, and an immunosuppressive tumor microenvironment (7). In addition, malignant CLL cells may provide suboptimal target-cell engagement because of altered expression of adhesion, costimulatory, and inhibitory molecules (7). Collectively, these features may impair CAR-T activation, expansion, persistence, and cytotoxic function despite preserved CD19 expression.

Combination strategies therefore have been explored to improve CAR-T activity in CLL. Ibrutinib has been evaluated as a partner for CD19 CAR-T therapy based on its ability to redistribute CLL cells from protective tissue niches, modulate tumor-cell interactions with the microenvironment, and improve T-cell fitness in patients with CLL (8,9). However, the optimal timing and contribution of BTKi exposure in the CAR-T setting remain incompletely defined, supporting investigation of additional immune-modulating approaches.

Lenalidomide is an immunomodulatory agent with established biological activity in CLL and related B-cell malignancies. Previous studies have shown that lenalidomide can modulate both T cells and malignant B cells, including effects on T-cell activation, cytokine production, costimulatory pathways, and tumor-cell susceptibility to immune-mediated killing (10). These properties make lenalidomide a rational candidate for combination with CD19 CAR-T therapy, particularly in CLL, where CAR-T efficacy may be constrained by both effector-cell dysfunction and inefficient target-cell engagement.

We therefore investigated lenalidomide as a combinatorial partner for CD19 CAR-T therapy in CLL. In parallel mechanistic studies, we evaluated the effects of lenalidomide on CAR-T phenotype, proliferation, cytotoxicity, cytokine secretion, transcriptomic programs, and interactions with primary CLL target cells.

We also initiated VTB-CLL002, an open-label, non-randomized phase 1/2 clinical trial evaluating CD19 CAR-T cells combined with early lenalidomide exposure and MRD-guided lenalidomide-based maintenance or consolidation in patients with relapsed/refractory CLL/SLL (ClinicalTrials.gov identifier: NCT06762431). Here, we report the mechanistic rationale, safety, cellular kinetics, and early clinical activity of this lenalidomide-containing CAR-T strategy.

## Results

### VTB-CLL002 study design and patient characteristics

VTB-CLL002 is an open-label, non-randomized phase 1/2 study evaluating CD19 CAR-T cells combined with early lenalidomide exposure and MRD-guided lenalidomide-based maintenance or consolidation in patients with relapsed/refractory CLL/SLL. Patients received ibrutinib before leukapheresis, obinutuzumab before cell collection to reduce circulating CLL burden and improve leukapheresis product composition, fludarabine/cyclophosphamide lymphodepletion, a single infusion of autologous CD19 CAR-T cells, and lenalidomide 10 mg orally from day 0 to day +6. Patients with MRD-positive disease after CAR-T therapy proceeded to lenalidomide plus obinutuzumab consolidation, whereas MRD-negative patients received lenalidomide maintenance **(Fig. 1)**.

**Figure 1.**
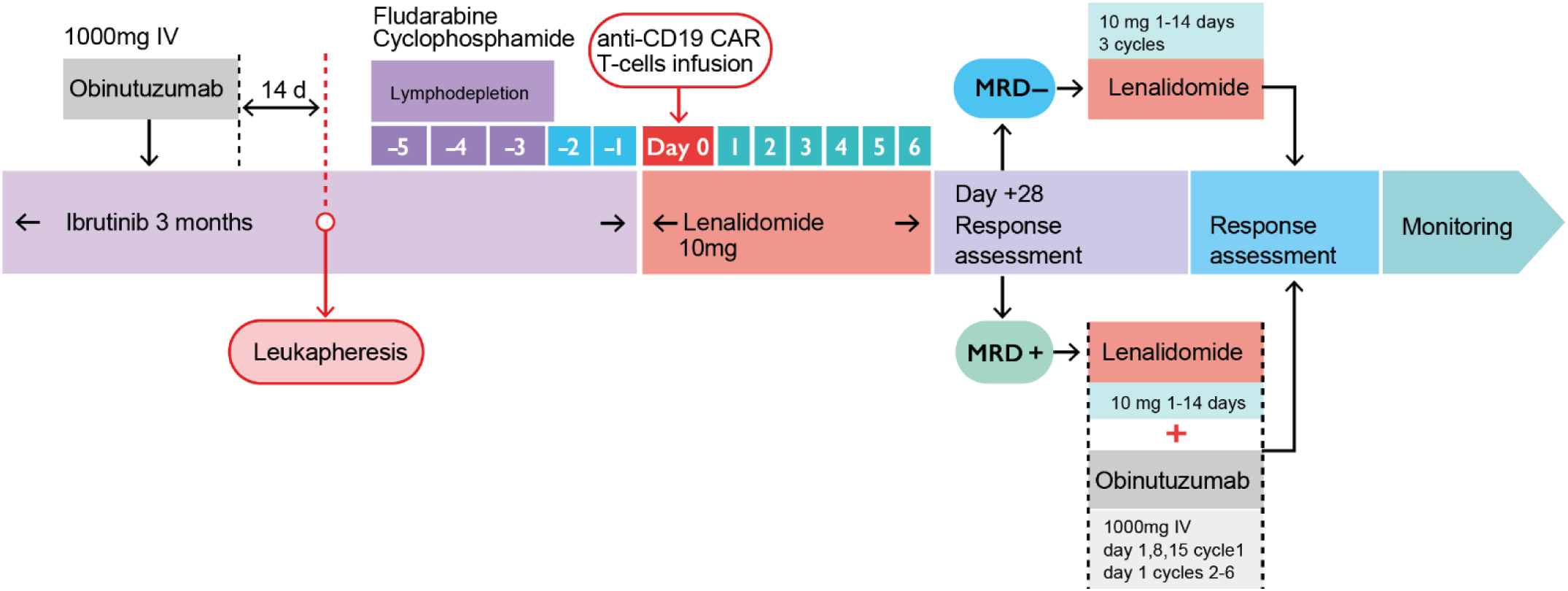
Flowchart of VTB-CLL002 clinical trial.

Between February 2024 and March 2026, 12 patients with relapsed/refractory CLL were screened and treated. The median age was 60 years (range, 46–67), and patients had received a median of 2 prior lines of therapy (range, 1–6). All patients were BTK inhibitor-naïve and had measurable disease at the time of CAR-T infusion. Most patients had unmutated IGHV status, and two patients had TP53 abnormalities. Baseline disease features included elevated LDH in 6 patients and bulky disease greater than 5 cm in 4 patients (Table 1). The median follow-up was 14 months (range, 1–24).

**Table 1.** Baseline characteristics of patients.

| Dose level | Patient ID | Gender | Age | Time from diagnosis | Prior therapy | TP53 aberration | IGHV mutation status | LDH>ULN | Bulky >5 cm | CAR-hematotox |
| --- | --- | --- | --- | --- | --- | --- | --- | --- | --- | --- |
| 1 | 1 | F | 46-50 | 8 | FCR, BR, GB, GL, VenR | - | UM | No | No | 2 |
| 1 | 2 | F | 46-50 | 8 | FCR, BR | + | UM | Yes | No | 2 |
| 1 | 3 | M | 46-50 | 3 | FCR | - | UM | Yes | No | 1 |
| 2 | 4 | M | 56-60 | 17 | CCR, BR | - | M | No | No | 2 |
| 2 | 5 | M | 61-65 | 3 | BR | - | UM | No | Yes | 1 |
| 2 | 6 | M | 56-60 | 7 | FCR, BR, R-BAC, R2, OFAR, FCM | + | UM | Yes | No | 2 |
| 3 | 7 | F | 56-60 | 2 | BR, R-CHOP, R-DHAP | - | N/A | Yes | Yes | 4 |
| 3 | 8 | F | 46-50 | 9 | FCR, BR | - | UM | No | Yes | 0 |
| 3 | 9 | M | 61-65 | 5 | FCR | - | UM | Yes | No | 2 |
| 3 | 10 | F | 61-65 | 10 | FCR, BR | - | UM | No | Yes | 2 |
| 3 | 11 | M | 61-65 | 4 | FCR | - | N/A | No | No | 1 |
| 3 | 12 | F | 61-65 | 8 | BR | - | UM | Yes | No | 0 |

LDH>ULN – Lactate Dehydrogenase > Upper Limit of Normal; F – female, M – male; IGHV - Immunoglobulin Heavy Chain Variable Region; M – mutated IGHV; UM- unmutated IGHV; N/A - not available; FCR - Fludarabine, Cyclophosphamide, Rituximab; BR - Bendamustine, Rituximab; GB - Obinutuzumab, Bendamustine; VenR - Venetoclax, Rituximab; CCR - Cladribine, Cyclophosphamide, Rituximab; R-BAC - Rituximab, Bendamustine, Cytarabine; R2 - Rituximab, Lenalidomide; R-CHOP - Rituximab, Cyclophosphamide, Doxorubicin, Prednisolone, Vincristine; OFAR - Rituximab, Fludarabine, Oxaliplatin, Cytarabine; FCM - Rituximab, Cyclophosphamide, Mitoxantrone, R-DHAP - Rituximab, Dexamethasone, Cytarabine, Cisplatin;

All patients received one CD19 CAR-T infusion across three dose levels: 25×10⁶ CAR-T cells in dose level 1, 50×10⁶ CAR-T cells in dose level 2, and 100×10⁶ CAR-T cells in dose level 3. Three patients with MRD-positive disease after initial response assessment received lenalidomide plus obinutuzumab consolidation, while the remaining patients received lenalidomide maintenance **(Fig. 1**; **Table 1)**.

### CD19 CAR-T manufacturing was feasible across all treated patients

CD19 CAR-T cell manufacturing was successful for all 12 treated patients. Final products were generated from autologous T cells transduced with a second-generation CD19 CAR construct and expanded ex vivo before fresh infusion. Despite marked heterogeneity in starting material and final product composition, all products met release criteria for clinical administration **(Table 2)**.

Characteristics and cellular composition of CD19 CAR T-cell products are summarized in Table 2. The median proportion of naïve and stem cell memory T cells (T_N_+T_SCM_) among CD4+ T cells was 3.3% (interquartile range [IQR]: 0.6–9.0%), and among CD8+ T cells — 3.4% (IQR: 0.4–5.3%). The median frequencies of CD4+ and CD8+ cells within CD3+CAR+ cells reached 44.9% (IQR: 35.8–71.1%) and 47.3% (IQR: 26.0–57.6%), respectively. *In vitro* proliferative capacity, assessed by total cell yield in the final product, exhibited a median of 2.94×10⁸ cells (range: 8.84×10⁷ – 4.00×10⁹). Median cell viability was 94% (range: 76–100%). The CD4/CD8 ratio of CAR-T cells ranged from 0.59 to 9.8, with a median of 2.1 (IQR: 0.94–4.66).

CAR transduction efficiency, defined as the percentage of CD3+CAR+ T cells, was also variable: median 26.5% (IQR: 14.5–34.3%). Two patients demonstrated low transduction efficiency — 4% and 6% CD3+CAR+ cells — despite a high total cell count in the products (7.65×10⁸ and 4.0×10⁹ cells, respectively). Minor populations of CD4-CD8- and CD4+CD8+ cells among CD3+ T lymphocytes did not exceed 5% and 3%, respectively, in the majority of products. Regarding differentiation phenotype, effector memory T cells (T_EM_) predominated within the CD8+ compartment (median 82.3%), whereas CD4+ cells displayed greater diversity: in individual patients, a substantial pool of T_N_+T_SCM_ was preserved (up to 37% of CD4+ T cells and 63% of CD8+ T-cells in patient Pt01) (Table 2).

### Combined lenalidomide and CD19 CAR-T therapy was associated with clinically meaningful toxicity

All treated patients experienced at least one adverse event of special interest. CRS occurred in all 12 patients, with severe CRS of grade ≥3 observed in 2 patients (16.7%). ICANS occurred in 5 patients (41.7%), including severe ICANS of grade ≥3 in 4 patients (33.3%). Three patients developed IEC-HS, all of which resolved with treatment. The maximum tolerated dose was not reached **(Table 3)**.

**Table 3.** Safety of all treated patients under VTB002 clinical trial. AE of special interest.

| ID | CRS | ICANS | IEC-HS | Infections | ICAHT |  | Other |
| --- | --- | --- | --- | --- | --- | --- | --- |
|  |  |  |  |  | Early | Late |  |
| 1 | 3 | - | + | Herpes 6 viremia, Stomatitis | 3 | 3 | HG, DIC |
| 2 | 1 | - | - | - | 1 | 4 |  |
| 3 | 2 | - | - | - | 4 | 3 | Dermatitis |
| 4 | 2 | - | - | Stomatitis | 3 | 2 | HG, DIC |
| 5 | 2 | 3 | + | IPA, Cl. Difficile enterocolitis | 1 | 3 | HG, DIC |
| 6 | 3 | 4 | + | IPA | 3 | 3 | HG, DIC<br>SCLC |
| 7 | 2 | 3 | - | - | 4 | 2 | HG |
| 8 | 3 | 1<br>4 grade late<br>ICANS | - | Stomatitis, COVID-19, CMV viremia, Pan resistant K. pneumonia sepsis | 3 | 3 | DIC<br>Bullous pemphigoid |
| 9 | 2 | - | - | - | - | 3 | DIC<br>Pulmonary embolism |
| 10 | 2 | - | - | - | 3 | 3 | DIC |
| 11 | 1 | - | - | - | 1 | 2 | DIC |
| 12 | 1 | 2 | - | - | 3 | 3 | DIC |

AE – adverse event; CRS – cytokine release syndrome, IEC-HS - Immune Effector Cell-Associated Hemophagocytic Lymphohistiocytosis-Like Syndrome, HG – Hypogammaglobulinemia (IgG<4g/L); DIC - Disseminated intravascular coagulation; IPA – Invasive pulmonary aspergillosis; CMV – Cytomegalovirus; SCLC – Small Cell Lung Cancer;

Hematologic toxicity was common. Early ICAHT occurred in 11 patients, including grade ≥3 events in 8 patients. Late ICAHT was observed in all patients, with grade ≥3 late ICAHT in 9 of 12 patients. Moderate or severe hypogammaglobulinemia occurred in 5 patients and was managed with intravenous immunoglobulin replacement. Transient disseminated intravascular coagulation was observed in 9 patients and was managed with fibrinogen concentrate, cryoprecipitate, and coagulation factor replacement as clinically indicated **(Table 3)**. Infectious complications included human herpesvirus 6 reactivation, cytomegalovirus reactivation, invasive pulmonary aspergillosis in two patients, Cl. difficile enterocolitis, COVID-19, and pan-resistant Klebsiella pneumoniae sepsis. One patient developed a secondary malignancy, small cell lung cancer, and one patient developed pulmonary embolism.

A notable safety event occurred in Patient 8. This patient initially developed grade 1 ICANS during the early post-infusion period, which was successfully managed. After reintroduction of lenalidomide more than 30 days after CAR-T infusion, the patient developed late-onset grade 4 ICANS requiring intensive care. This episode was temporally associated with secondary CAR-T expansion and was complicated by COVID-19 and fatal pan-resistant *K. pneumoniae sepsis* **(Table 3**; **Table 4)**. Although causality cannot be established in this single case, the observation suggests that post-infusion lenalidomide exposure may contribute to delayed CAR-T reactivation in selected patients and should be evaluated carefully in future dosing schedules.

**Table 4.** Cellular kinetics, efficacy and monitoring of treated patients.

| Patient ID | Cmax (cells/μL) | CD4/CD8 at peak expansion | Tmax (days) | Consolidation* | Best response (months) | MRD best response (days) | Follow up (months) | CAR T-cell persistence at last Follow up |
| --- | --- | --- | --- | --- | --- | --- | --- | --- |
| 1 | 48 | 0,01 | +18 | - | CR (+6) | MRD - (+18) | 16 | yes |
| 2 | 51 | 0,98 | +21 | - | CR (+1) | MRD - (+30) | 15 | yes |
| 3 | 10,42/<br>73,34 | 0,9/0,38 | +19/+60 | + | CR (+1) | MRD - (+60) | 15 | yes |
| 4 | 27,9 | 4,7 | +15 | - | CR (+1) | MRD - (+5) | 13 | yes |
| 5 | 524 | 0,02 | +9 | + | CR (+3) | MRD - (+21) | 21 | yes |
| 6 | 369 | 0,14 | +17 | - | CR (+1) | MRD - (+28) | 24 | yes |
| 7 | 35,9 | 1,43 | +7 | - | CR (+1) | MRD - (+28) | 16 | yes |
| 8 | 5995 | 0,03 | +18 | - | CR (+1) | MRD - (0) | 8 | yes |
| 9 | 25,35 | 0,8 | +13 | + | CR (+1) | MRD - (+62) | 3 | yes |
| 10 | 211 | 0,5 | +13 | - | CR (+1) | MRD - (+6) | 3 | yes |
| 11 | 473 | 0,34 | +9 | - | CR (+1) | MRD - (0) | 2 | yes |
| 12 | 346 | 0,12 | +8 | - | CR (+1) | MRD - (0) | 1 | yes |

CR – complete response; MRD – minimal residual disease; Cmax - peak concentration of CAR T-cells; Tmax – time to peak concentration of CAR T-cells; \* Patients with MRD+ disease after CD19 CAR T-cell therapy receiving lenalidomide plus obinutuzumab consolidation

Overall, the regimen was feasible, but toxicity was clinically meaningful, particularly with respect to neurotoxicity and hematotoxicity. These findings support continued clinical evaluation with careful attention to lenalidomide timing, post-infusion re-exposure, and monitoring for delayed immune-effector toxicity.

### CAR-T expansion, persistence, and clinical responses

*In vivo* expansion of CD19 CAR T-cells was observed in all treated patients, with a median peak expansion of 137 cells/µL (range: 10.42 - 5995 cells/µL) and a median time to peak expansion (Tmax) of 14 days (range: 7 – 60 days). Although the majority of the infused CAR T-cell products were CD4-dominant **(Table 2)**, a shift toward CD8 dominance was observed in 10 of 12 patients (83.3%) at the time of peak expansion **(Table 4)**.

In Patient 3, who remained MRD-positive at day +30, CAR T-cell counts significantly increased from 3 cells/µL to 73.34 cells/µL following consolidation therapy with lenalidomide and obinutuzumab. This secondary expansion suggests potential cytokine-mediated support and reinvigoration of the CAR T-cell population.

Notably, CD19 CAR T-cells remained detectable by flow cytometry in all patients at the last follow-up, with persistence documented for up to 2 years. The maximum peak expansion (5995 cells/µL) was recorded in Patient 8; following lenalidomide consolidation, this patient experienced a secondary wave of expansion that correlated with the development of late-onset ICANS.

All patients were evaluable for response. Within the DL1 cohort, Patient 1 received a product with the highest proportion of memory CAR T-cells (63% of Tn/Tscm CD8+ and 37% of Tn/Tscm CD4+ T-cells; **Table 2**). This patient achieved an MRD-negative CR that has been sustained for over 20 months, distinguishing this case from others in the DL1 group. This observation is consistent with previous reports suggesting that therapeutic doses can be reduced up to 25-fold when the CAR T-cell product is enriched with the Tscm subset (**11**), further underscoring the clinical value of Tscm enrichment. We may speculate that lenalidomide is responsible for the depth of response, while the CAR+ Tscm content ensures the durability of the response.

Overall, all patients achieved a CR with a median time to response of 1 month (range: 1–6 months), demonstrating a robust capacity for debulking even high tumor volumes **(Fig. 2A)**. Two patients from the DL1 cohort (Patient 2 and Patient 3) achieved an MRD-negative CR even at this low dose, though they eventually converted from MRD-negative to MRD-positive status **(Fig. 2B, 2C)**. In contrast, eight of nine patients from the DL2 and DL3 cohorts show an ongoing MRD-negative response with a median duration of 8 months (range: 1–24 months). Median PFS and OS have not yet been reached in this group **(Fig. 2B, 2C)**, although a longer observation period is required.

**Figure 2.**
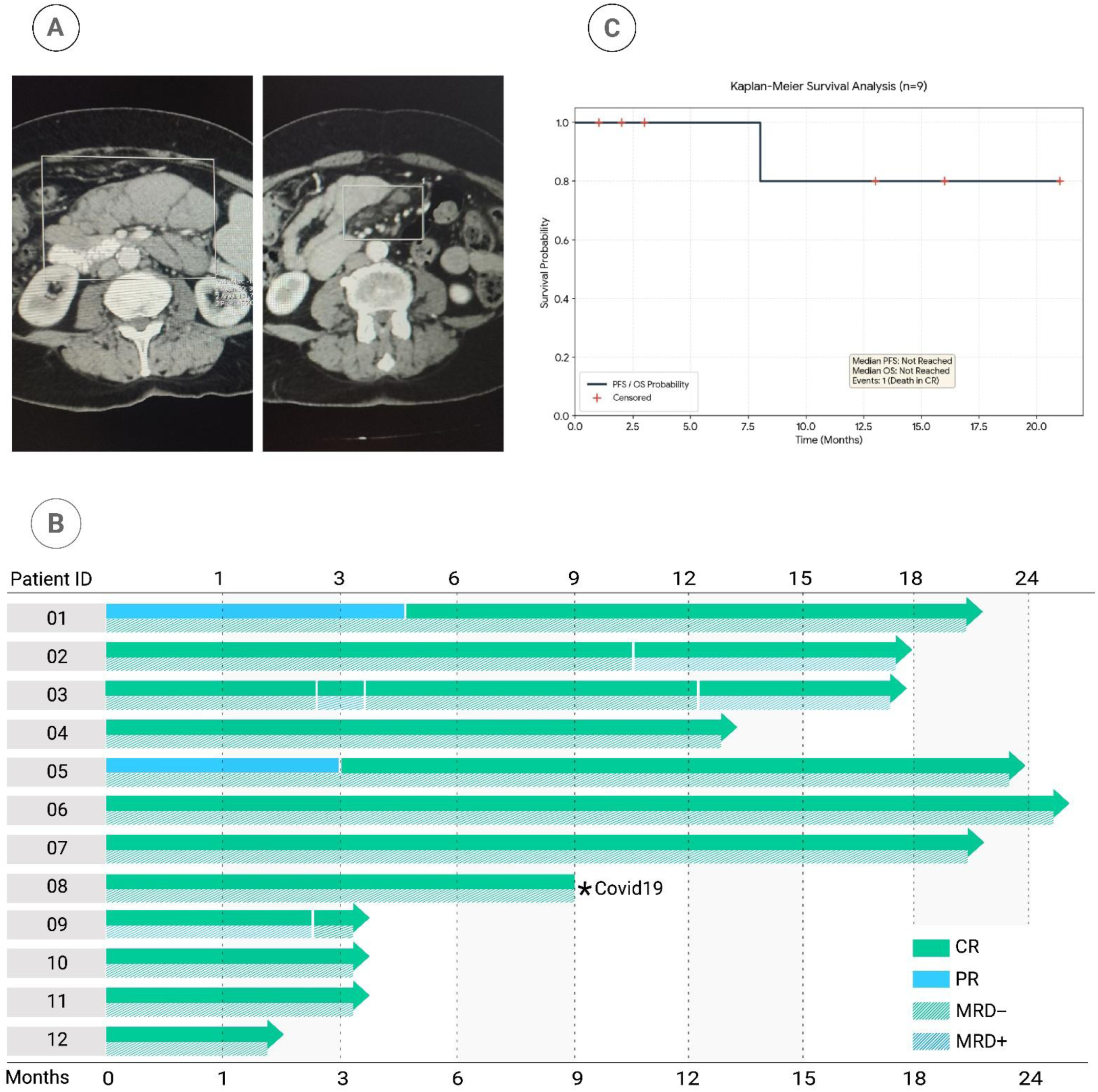
Efficacy of CD19 CAR T-cells with concomitant lenalidomide for R/R CLL in the VTB002 phase 1/2 clinical trial. A) Representative CT scans of Patient 10 showing high tumor burden before and after treatment with CD19 CAR T-cells and lenalidomide. B) Swimmer plot illustrating clinical outcomes and duration of response for all treated patients. Abbreviations: CR, complete response; PR, partial remission; MRD–, minimal residual disease-negative; MRD+, minimal residual disease-positive. C) Kaplan-Meier survival curves. Survival analysis was performed for the DL1 and DL2 cohort (n=9). One event (death during remission) occurred at month 8.

### Lenalidomide enhances CAR-T proliferation and induces transcriptomic remodeling of T-cell phenotype

The immunostimulatory impact of lenalidomide on T and NK cells— characterized by enhanced co-stimulation, regulatory T-cell suppression, and cereblon-mediated IKZF1/3 degradation (12–14). Furthermore, ibrutinib exerts distinct immunomodulatory effects on T and B cells through the inhibition of Bruton’s tyrosine kinase (BTK) and interleukin-2-inducible T-cell kinase (ITK), and is already widely employed to modulate CAR-T cell exhaustion and preserve their functional activity (15,16). Building on these complementary pathways, we investigate the underlying mechanisms of lenalidomide alone and in combination with ibrutinib on CAR-T cells, proposing this dual immunomodulation as a potentially promising strategy to synergistically enhance CAR-T fitness and persistence (**Fig. 3A**).

**Figure 3.**
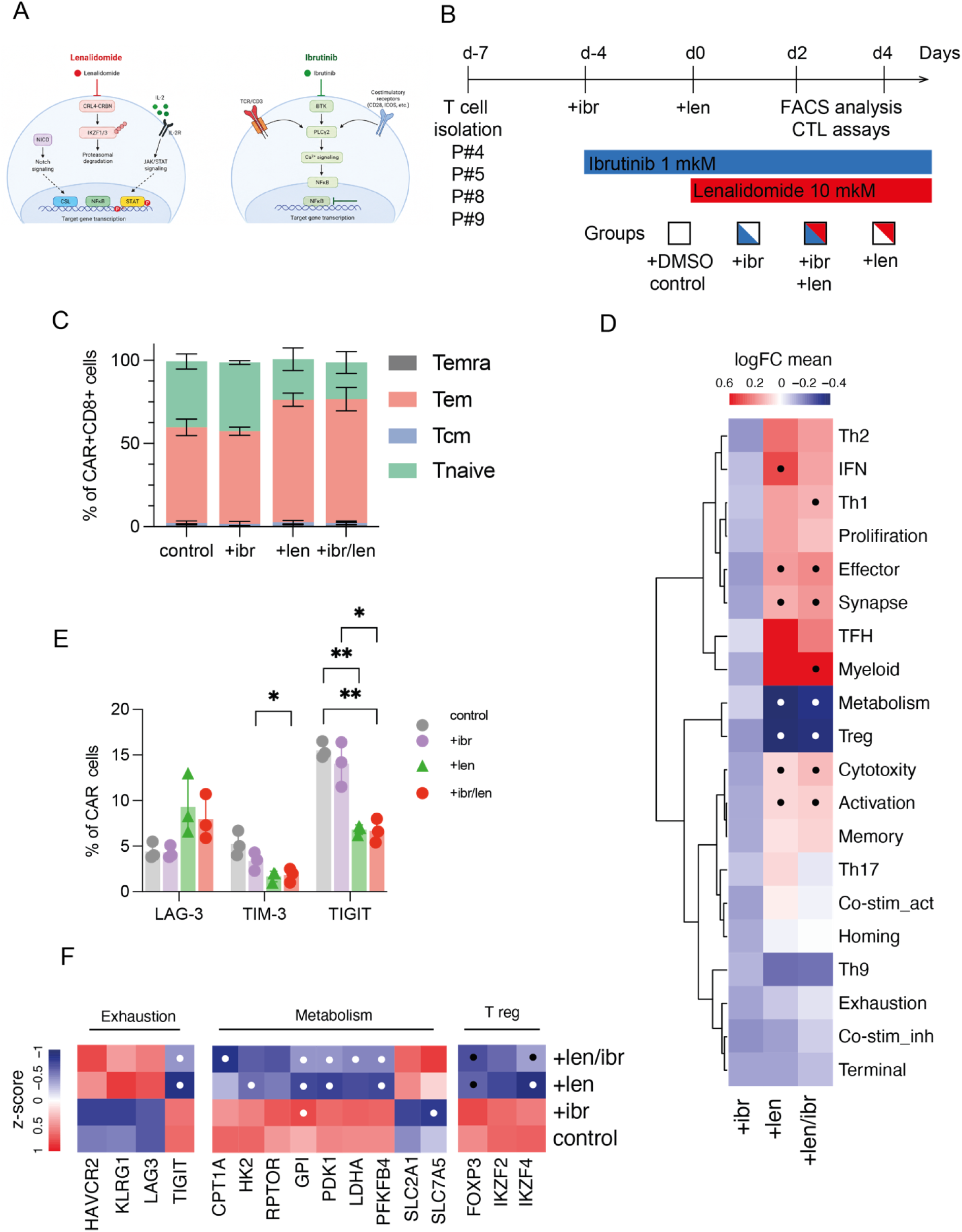
Lenalidomide enhances CAR-T proliferation and phenotype. **A.** Schematic representation of the molecular mechanisms underlying lenalidomide and ibrutinib activity. **B.** Experimental design of the in vitro CAR-T modulation assay. CD19 CAR-T cells were cultured under four conditions: vehicle control (DMSO), ibrutinib alone (1 µM), lenalidomide alone (10 µM), or combined ibrutinib and lenalidomide. Ibrutinib was added on day −4, and lenalidomide was added on day 0. **C.** Differentiation phenotype of CD4⁺ and CD8⁺ CAR-T cells on day 4, assessed by flow cytometry. **D.** Heatmap of pathway enrichment analysis for each treatment group relative to vehicle control after 24 hours of stimulation. **E.** Expression of exhaustion LAG-3, TIM-3, TIGIT markers on day 4, assessed by flow cytometry. **F.** Heatmap of exhaustion, metabolism and T-regulatory associated genes after 24 hours of stimulation. Data are shown as mean ± SD. A minimum of three independent donors was analyzed across panels. Statistical tests are indicated for each panel: **E-F** - one-way ANOVA followed by Tukey’s multiple-comparison test, as appropriate. Only significant p values are shown.

To determine whether lenalidomide directly modulates CAR-T cell function, CD19 CAR-T cells were generated from healthy donors and patients and cultured under four conditions: vehicle control, ibrutinib, lenalidomide, or combined ibrutinib and lenalidomide. Ibrutinib was introduced before CAR-T stimulation to model pre-exposure, whereas lenalidomide was added at the beginning of the functional assays (**Fig. 3B**). Lenalidomide increased CAR-T cell proliferation compared with vehicle-treated controls (**Supplementary Fig. 2A**). Early after exposure, exhaustion marker expression was not substantially altered (**Supplementary Fig. 2B**). However, by day 4, lenalidomide-containing conditions were associated with a shift toward effector-associated phenotypes, including enrichment of effector memory CD8^+^ CAR-T cells (p-value = 0.0075) and reduction in len+ of naïve T-cell populations in comparison with control group (CD4^+^: p-value = 0.0169, CD8^+^ p-value = 0.0323) (**Fig. 3C**).

To investigate transcriptional correlates of lenalidomide-enhanced CAR-T function, sorted CAR⁺ T cells exposed to lenalidomide, ibrutinib, or the combination were analyzed using NanoString gene expression profiling (**Supplement Fig. 3A,B**). Transcriptomic profiles of ibrutinib-treated and control CAR-T cells were relatively similar (**Supplement Fig. 3A)**. In contrast, lenalidomide-containing conditions induced broader transcriptional reprogramming, with approximately 55 genes upregulated and 14–18 genes downregulated compared with control conditions (**Supplement Fig. 3 B-D**). The RNA-seq analysis corroborated the phenotypic shift and revealed a significant upregulation of gene signatures associated with T-cell activation and the effector phenotype (**Fig. 3D, Supplement Fig. 3 C,D**). Combined ibrutinib and lenalidomide exposure was associated with reduced expression of selected exhaustion markers, including TIGIT and TIM-3, whereas effects on LAG-3 were more variable (**Fig. 3D, F**). This reduction in TIGIT expression was further validated at the transcriptional level by RNA-seq, which confirmed decreased *TIGIT* mRNA levels while other exhaustion-associated genes remained unchanged, suggesting selective modulation of the exhaustion phenotype. (**Fig. 3F**). Beyond these transcriptional changes, we also observed profound effects of Lenalidomide on cellular metabolism. Metabolic profiling revealed a distinct reprogramming pattern in lenalidomide-containing conditions, characterized by downregulation of oxidative phosphorylation and glycolytic pathway genes (*CPT1A, HK2, mTOR, PDK1, LDHA, PFKFB4*), concomitant with upregulation of nutrient transporters *SLC2A1 (GLUT1)* and *SLC7A5 (LAT1)*, suggesting a metabolic shift toward enhanced nutrient uptake capacity with reduced metabolic flux **(Fig. 3F).** Furthermore, transcriptomic analysis revealed a suppression of the regulatory T-cell (Treg) program, evidenced by the downregulation of FOXP3, IKZF2, and IKZF4 **(Fig. 3F).** This transcriptional signature aligns with the moderate reduction in Treg cell populations observed in patients following lenalidomide therapy. **(Supplementary Fig. 2B)**

In summary, lenalidomide comprehensively reshapes CAR-T cells into a highly activated effector state and primes them for enhanced cytotoxicity and target engagement by rewiring their metabolic and transcriptional networks (**Fig. 3, Supplementary Fig. 2-3).**

### Lenalidomide enhances CAR-T effector function and remodels immune synapse formation

We next assessed whether these phenotypic changes translated into improved antitumor activity of CAR-T cells. In short-term cytotoxicity assays against CD19-positive C-II targets, lenalidomide-treated CAR-T cells showed enhanced target-cell killing compared with vehicle-treated CAR-T cells (**Fig. 4A**). Combined lenalidomide and ibrutinib exposure further increased cytotoxicity in several assay conditions compared with ibrutinib alone, supporting an additive functional effect rather than a purely phenotypic change (**Fig. 4A**).

**Figure 4.**
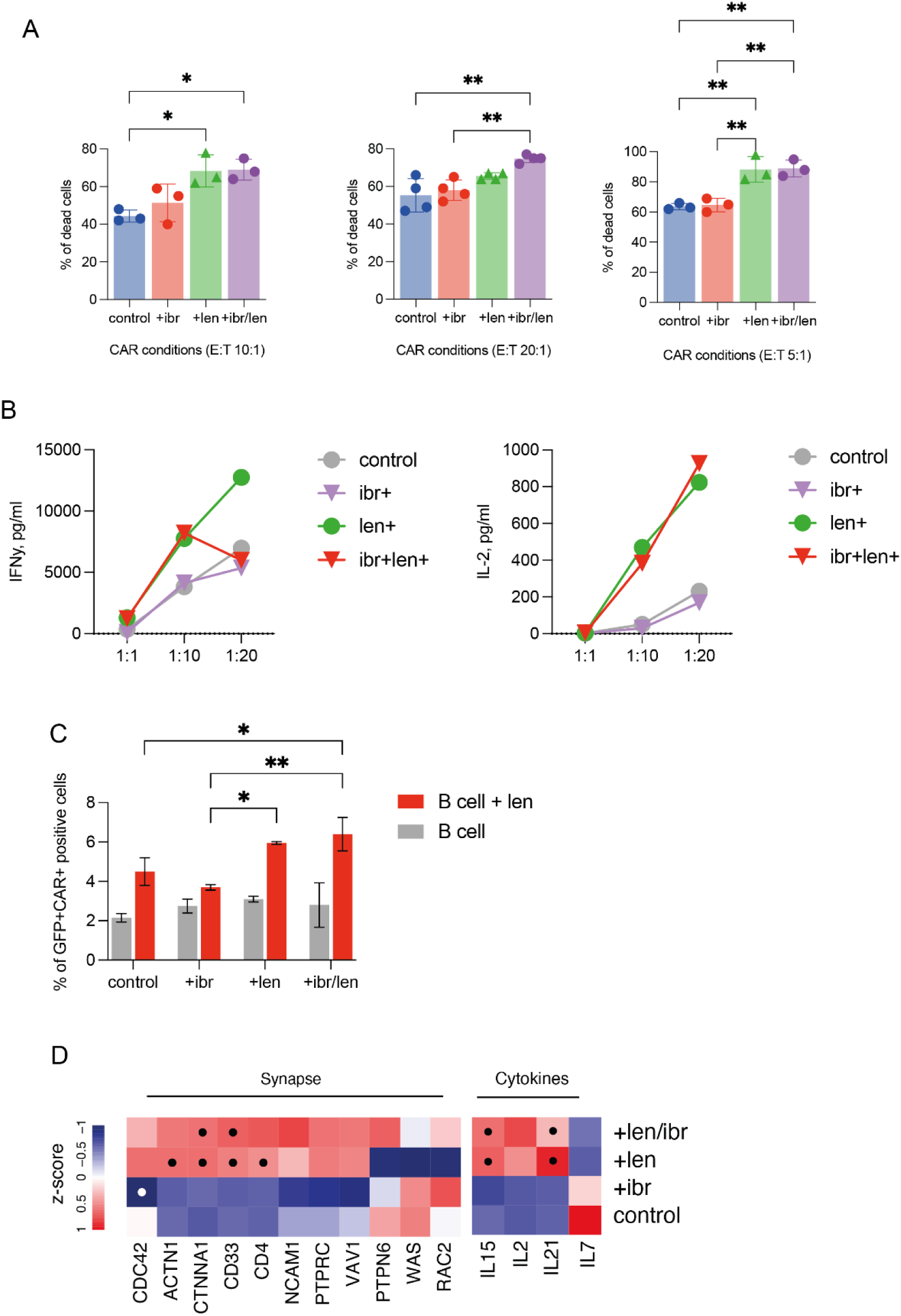
Lenalidomide enhances CAR-T cytotoxicity and cytokine secretion. **A.** Cytotoxic activity of CD19 CAR-T cells cultured under the indicated treatment conditions against CII target cells at different effector-to-target ratios. **B.** IL-2 and IFN-γ secretion by autologous CD19 CAR-T cells after 24-hour co-culture with patient-derived CLL cells. Non-transduced T cells were included as a negative control. **C.** Quantification of CAR-T/CLL conjugate formation after co-culture. Cells were stained for CD19 and CAR/EGFRt, and double-positive events were quantified as CAR-T/CLL conjugates. This flow-based assay was used as a measure of target-cell engagement. **D.** Heatmap of selected genes associated with T-cell cytokine signaling and synapse formation across treatment groups relative to vehicle control. P-values are shown with dot. Data are shown as mean ± SD. A minimum of three independent donors was analyzed across panels. Statistical tests are indicated for each panel: **A,D** - one-way ANOVA followed by Tukey’s multiple-comparison test, **C** - repeated-measures two-way ANOVA with Geisser–Greenhouse correction followed by Sidak’s or Tukey’s multiple-comparison test, as appropriate. Only significant p values are shown.

Consistent with improved productive engagement, lenalidomide-treated CAR-T cells secreted higher levels of IL-2 and IFN-γ after co-culture with CD19-positive CLL targets (**Fig. 4B**). This enhanced cytokine production was accompanied by upregulation of *IL15* and *IL21* genes in CAR-T cells (**Fig. 4С**).

To model repeated antigen exposure, CAR-T cells were evaluated in prolonged co-culture and rechallenge assays (**Supplement Fig. 4A).** Under these conditions, lenalidomide-treated CAR-T cells maintained more favorable functional activity during serial tumor challenge and showed reduced frequencies of TIGIT⁺, TIM-3⁺, and LAG-3⁺ CAR-T cells compared with controls (**Supplement Fig. 4B–C**). These findings suggest that lenalidomide can preserve CAR-T function during sustained antigen stimulation, a setting relevant to CLL where persistent tumor exposure may contribute to CAR-T dysfunction.

To evaluate CAR-T/CLL target-cell engagement, co-cultures were stained for CAR and CD19, and double-positive events were quantified as CAR-T/CLL conjugates. Lenalidomide increased the frequency of CAR⁺CD19⁺ conjugates, particularly when both CAR-T cells and CLL cells were exposed to lenalidomide (**Fig. 4D**). Moreover, transcriptomic profiling of CAR-T cells revealed significant upregulation of key synapse-associated genes, including *CDC42, ACTN1, CTNNA1, CD33*, and *CD4*, which are critical for cytoskeletal reorganization and adhesion molecule clustering at the immunological synapse (Fig. 5H) (17). Additionally, genes regulating signal transduction at the synapse, such as *VAV1, PTPN6*, and *WAS*, showed increased expression in lenalidomide-treated conditions (**Fig. 4C**). These transcriptional changes suggest that lenalidomide promotes the molecular machinery necessary for stable CAR-T/CLL cell conjugate formation and effective immune synapse assembly.

**Figure 5.**
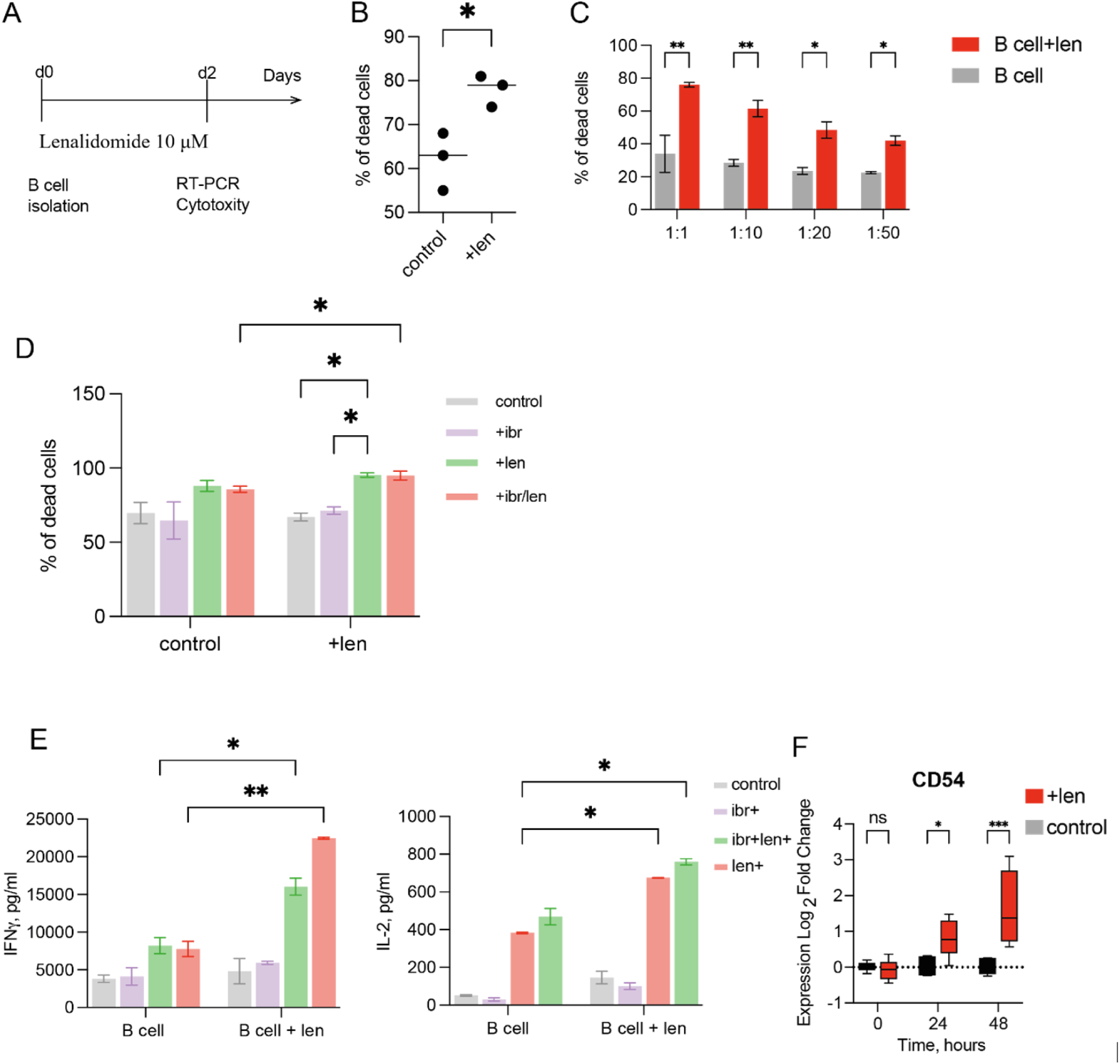
Lenalidomide improves CAR-T activity against primary CLL targets. **A.** Experimental design of the in vitro CLL modulation assay. CLL cells were cultured under two conditions: vehicle control (DMSO) and lenalidomide alone (10 µM). **B.** Cytotoxic activity of autologous CD19 CAR-T cells against CLL cells (C-II) pretreated with lenalidomide or vehicle control. CLL target cells were exposed to lenalidomide, washed, and then co-cultured with CAR-T cells at 1:5 effector-to-target ratios. **C**. Cytotoxic activity of CD19 CAR-T cells cultured under the indicated treatment conditions against lenalidomide-pretreated CLL cells or vehicle control. **D.** Cytotoxic activity of CD19 CAR-T cells cultured under the indicated treatment conditions against lenalidomide-pretreated patient-derived CLL cells pretreated with lenalidomide or vehicle control. CLL target cells were exposed to lenalidomide, washed, and then co-cultured with CAR-T. **E.** IL-2 and IFN-γ secretion by autologous CD19 CAR-T cells after 24-hour co-culture with patient-derived CLL cells. CLL cells were treated with lenalidomide or vehicle control before co-culture. Non-transduced T cells were included as a negative control. **F.** RT-qPCR analysis of CD54/ICAM-1 expression in patient-derived CLL cells cultured with lenalidomide or vehicle control for 0, 24, or 48 hours. Three independent donors were analyzed across panels. Data are shown as mean ± SD. Each dot represents an independent donor. Statistical analysis was performed using paired t-test for **B**, one-way ANOVA followed by Tukey’s multiple-comparison test for panel **C**, two-way ANOVA with Geisser–Greenhouse correction followed by Sidak’s multiple-comparison test for panel **D, E,F**. Only significant p values are shown.

Together, these data indicate that lenalidomide and combination of lenalidomide and ibrutinib enhances CAR-T proliferation and cytotoxicity while supporting a less exhausted, effector-associated phenotype during repeated antigen challenge (**Fig. 4; Supplement Fig. 4**). The transcriptional findings support the functional observation that lenalidomide enhances CAR-T effector activity and promotes a more activated, cytotoxic, and target-engagement-competent state. Together, the phenotypic, functional, target-cell, and transcriptomic data provide mechanistic correlates for the clinical activity observed in VTB-CLL002 (**Fig. 3–4**).

### Lenalidomide sensitizes primary CLL targets to CAR-T cell-mediated cytotoxicity

We next sought to determine whether lenalidomide’s effects extend beyond CAR-T cell modulation to include direct activity against primary malignant B cells from patients with CLL. To assess direct effects on tumor cells, we pre-incubated primary CLL target cells with lenalidomide, followed by thorough washing before co-culture with CAR-T cells (**Fig. 5A**). We observed increased CAR-T-mediated killing of lenalidomide-pre-treated CLL cells compared with vehicle-treated controls, indicating that lenalidomide directly modifies the susceptibility of malignant B cells to CAR-T attack (**Fig. 5B,C**). In these assays, target-cell lenalidomide exposure increased cytotoxicity from approximately 55% in control conditions to approximately 80% in the lenalidomide-treated condition (**Fig. 5C**).

The strongest cytotoxicity and IL-2 and IFN-γ secretion was observed when both compartments were exposed to lenalidomide: CAR-T cells treated with lenalidomide and CLL cells pretreated with lenalidomide showed higher cytokines secretion than either compartment alone (**Fig. 5D,E; Supplementary Fig. 5**). These findings support a dual-compartment mechanism in which lenalidomide improves both effector-cell function and target-cell susceptibility.

To define how lenalidomide modifies malignant B-cell susceptibility to CAR-T engagement, primary CLL cells were incubated with lenalidomide and assessed for changes in surface phenotype and gene expression. Lenalidomide did not broadly induce classical costimulatory molecules, including CD80, CD86, or CD40, by flow cytometry or RT-qPCR (**Supplementary Fig. 6**). While, CD58/LFA-3 expression was reduced (**Supplementary Fig. 5**). In contrast, Lenalidomide increased expression of CD54/ICAM-1, an adhesion molecule involved in T-cell target-cell interactions (**Fig. 5F**).

Together, these data establish that lenalidomide directly enhances the susceptibility of primary CLL cells to CAR-T attack through selective upregulation of CD54/ICAM-1, independent of classical costimulatory pathways. This target-cell sensitization, combined with lenalidomide’s parallel enhancement of CAR-T effector function, creates a dual-compartment mechanism that maximizes cytotoxicity and cytokine secretion, thereby explaining the superior anti-tumor efficacy when both compartments are exposed to lenalidomide (**Fig. 5; Supplementary Fig. 6**).

## Discussion

The approval of Lisocabtagene maraleucel (liso-cel) was based on the TRANSCEND CLL–004 phase 1/2 clinical trial, in which 118 patients were enrolled, and 97 of these were evaluable for efficacy (5,18). An overall response was achieved in 44% of patients, with 20% attaining complete remission and 64% achieving undetectable minimal residual disease (uMRD) negativity in blood. The median duration of response was 35.3 months, while the median duration of CR/CRi was not reached; however, the median PFS was only 2.8 months in patients with detectable MRD, underscoring the need for further strategies to improve treatment outcomes (5,19). The clinical efficacy of CD19 CAR T-cell therapy in CLL is at least two times lower compared to those observed in other B-cell malignancies following treatment (3). The inability of T-cells to form effective immunologic synapses represents a widespread immune dysfunction in CLL and is responsible for decreased activity of CAR T-cells in this connection. Several mechanisms have been proposed to explain this phenomenon such as hyperexpression of CD200, CD270, CD274, CD276 (10), CD24 and CD52 (20) with decreased expression of adhesion and co-stimulatory molecules on CLL cells. Collectively these alterations result in reduced activation and killing capacity of CAR T-cells against CLL cells (21).

Due to the suboptimal results demonstrated by CD19 CAR T-cells in CLL alone, a combination strategies incorporating ibrutinib to CD19 CAR T-cell therapy have been tested, relying on its ability to downregulate CXCR4 and other adhesion molecules such as CD49d on CLL cells (8,22). This typically results in rapid redistribution of CLL cells from spleen and lymph nodes into the circulation and their detachment from the immunosuppressive tumor microenvironment (23), potentially increasing their accessibility to circulating CAR T-cells (24). Additionally, ibrutinib has been shown to significantly increase CD4+ and CD8+ T-cell counts, broaden the T-cell receptor (TCR) repertoire, and improve T-cell fitness in CLL patients after three months of therapy (25). It also reduces the expression of immunosuppressive molecules such as PD-1 and CTLA-4, promotes Th1 polarization, enhances T-cells with memory properties, and inhibits myeloid-derived suppressor cells (MDSCs) (26,27). Furthermore, ibrutinib decreases expression of CD200 and BTLA and reduces IL-10 production by CLL cells (28), contributing to improved immune synapse formation and collectively modulating the tumor microenvironment to potentially enhance CAR T-cell efficacy (29). Several Phase I/II clinical trials have investigated the combination of second-generation CD19 CAR T-cells and ibrutinib, demonstrating approximately twice the improvement in ORR and CR rate compared to patients who did not receive ibrutinib (5,30). However, Gauthier J. et al. compared their Phase I/II trial of CD19 CAR T-cells with concurrent ibrutinib to previous studies where patients received CD19 CAR T-cell therapy after pre-treatment with ibrutinib, and they observed similar PFS. These findings raise the question of whether concurrent administration or pre-treatment with ibrutinib—has the most significant impact on the efficacy of ibrutinib in the context of CD19 CAR T-cell therapy (31,32), leaning more towards pre-treatment being potentially more responsible for the improved outcome. One of the reasons explained could be the negative impact of ibrutinib on activation and effector functions of T-cells against CLL cells in vivo (33).

Patients with CLL exhibit immune defects similar to those observed in individuals with inherited CD154 deficiency (34). Specifically, CD4+ T-cells from CLL patients fail to express CD154 upon ligation of CD3 (35). A well-established approach to address this involves co-culturing CLL cells with CD40L-expressing feeder cells; CD40 signaling in this context has been shown to upregulate the expression of various costimulatory (CD80, CD86 etc.) and adhesion molecules (CD54, CD58) (36) on the surface of CLL cells, while downregulating inhibitory molecules such as CD24 and CD52 (20). This strategy has demonstrated the ability to break T-cell anergy, restore immunological synapse formation, enhance T-cell activation, and improve cytotoxic responses against CLL cells (20,21,37–41).

The immunomodulatory drug lenalidomide has been shown to repair autologous T-cell synapse formation defects in CLL by downregulating inhibitory ligands (such as CD200, CD270, CD274, and CD276) and their receptors on T cells (10). Additionally, lenalidomide induces the functional expression of CD154 on CLL cells, promoting a maturation process similar to that observed after trans CD40 ligation (42). Notably, CD40L expression has also been observed on CAR T-cells following lenalidomide exposure (43). In our experiments, an upregulation of CD80/CD86 and CD40 molecules was not detected. This could be attributed to experimental limitations; specifically, the CLL specimens were obtained after patient exposure to ibrutinib.

Numerous studies have demonstrated that CAR T-cells can enhance cancer cell killing capacity through mechanisms such as increased activation through pERK phosphorylation and cytokine production (IFNy, IL-2 etc.) (44), proliferation, persistence, tumor infiltration, and modulation of the immunosuppressive tumor microenvironment and Th1 CD4 T-cells polarization, depletion of Treg T-cells and MDSC (43–48). Furthermore, early exposure to lenalidomide at the time of CD19 CAR T-cell expansion has been associated with high response rates in very high-risk patients experiencing relapse after CAR T-cell therapy for relapsed/refractory aggressive large B-cell lymphoma (49). Our *in vitro* data demonstrate that CLL cells pre-exposed to ibrutinib and subsequently treated with lenalidomide exhibit a clear upregulation of CD54 (ICAM-1) molecules. This upregulation promotes adhesion and immunological synapse formation with CD19 CAR T-cells. Consequently, in co-culture models, this combination enhances the production of IL-2, IL-15, IL-21, and IFNy cytokines, skews T-cells toward a Th1 and effector phenotype, and increases the secretion of granzyme B and perforin, thereby boosting overall cytotoxicity. Previous studies have demonstrated a critical role for CTLA-4 in suppressing CD19 CAR T-cell proliferation and the cytotoxic capacity of CLL cells; notably, the knockout of CTLA-4 significantly enhanced the performance of CD19 CAR T-cells against CLL (50). Our data demonstrate no changes in the CD80 and CD86 CTLA-4 ligands on CLL cells. This phenotype potentially diminishes CTLA-4 signaling on CD19 CAR T-cells, thereby contributing to the enhanced efficacy of lenalidomide in this context.

While the preliminary clinical outcomes in our study compares favourably those reported for both liso-cel monotherapy (5) and the liso-cel plus ibrutinib combination (51) at the same stage of investigation, the current median observation time is too short to definitively determine if the addition of lenalidomide enhances response durability and PFS. Although our clinical trial demonstrated a seemingly higher incidence of high-grade ICANS compared to CD19 CAR T-cell therapy (18), all adverse events, including ICANS, remained manageable. Nevertheless, implementing additional prophylactic and therapeutic strategies will be essential for future trials.

The lack of a control group is a further limitation, as ibrutinib, obinutuzumab, and lenalidomide might have provided supplemental clinical benefit; however, given the brief duration of exposure, their impact on CR rates is likely negligible. Moreover, an additional limitation of the present study is the lack of a stratified analysis evaluating lenalidomide-induced activity within the CD19 CAR T-cell and CLL microenvironment across different molecular and cytogenetic risk groups. Given that specific genomic aberrations, such as del(17p), TP53 mutations, and NOTCH1 mutations, significantly influence both CAR T-cell fitness and tumor cell susceptibility, evaluating how these high-risk profiles modulate the therapeutic efficacy of lenalidomide remains a critical objective. Consequently, both future preclinical investigations and clinical trials involving larger patient cohorts are warranted to elucidate these complex interactions and optimize personalized therapeutic strategies.

## Methods

### Study design and CD19 CAR T-cells manufacturing

This is an open-label, non-randomized Phase 1/2 clinical trial, VTB-CLL002 (ClinicalTrials.gov identifier: NCT06762431), evaluating the safety and efficacy of CD19 CAR-T cell therapy combined with concomitant lenalidomide in patients with relapsed or refractory CLL and SLL, followed by MRD-guided consolidation (Figure 1). To be eligible, iBTK-naive patients with CLL needed to have failed at least one prior line of therapy and have an Eastern Cooperative Oncology Group (ECOG) performance status of 0 or 1. After signing informed consent, treatment with ibrutinib at the standard dose of 420 mg orally daily for three months up to day -1 was initiated. Two weeks prior to leukapheresis, patients received intravenous infusion of obinutuzumab 1000 mg to reduce circulating CLL cells and improve the processing of peripheral blood for ex vivo CAR T-cell preparation and get rid of an unfavorable T-cell:B-cell ratio in the leukapheresis products. Lymphodepletion consisted of fludarabine 25 mg/m^2^ and cyclophosphamide 250 mg/m^2^ for 3 consecutive days (from day-5 to day-3).

In a 3+3 clinical trial design, patients received increasing doses (from 25 to 100 million) of autologous T cells lentivirally transduced to express a second-generation CD19 CAR-T cell construct (FMC63.IgG4.BBz) as previously described (52). Fresh CD19 CAR T-cells were manufactured at the Biotechnology Laboratory at the Belarusian Research Center of Pediatric Oncology, Hematology, and Immunology and transferred to the Vitebsk regional clinical cancer Centre, where the infusion took place. Patients received lenalidomide 10 mg per os from day 0 to day +6. Response assessment was performed one month after infusion. Based on peripheral blood minimal residual disease (MRD) assessment by flow cytometry, MRD positive patients are suggested to undergo six cycles of combination therapy of lenalidomide and obinutuzumab, while MRD negative patients received 3 cycles of lenalidomide maintenance therapy. The trial was approved by the institutional review board as well as by the local regulatory authority. The study was conducted in accordance with the Declaration of Helsinki and International Conference on Harmonization guidelines for Good Clinical Practice.

### Flow cytometry

Flow cytometry acquisition was performed with a DxFlex flow cytometer (Beckman Coulter, C78505). CAR identification on the surface of CAR T cells was performed by biotinylated recombinant CD19 protein (in-house production) and streptavidin-APC conjugate (BioLegend, 405207). tEGFR reporter was identified by the Alexa Fluor 488-conjugated biosimilar of cetuximab (R&D Systems, FAB9577G-100). The phenotypic composition and quantity of T lymphocytes in the patient’s blood before apheresis, PBMCs, and final CAR T product were determined by staining the cells with antibodies CD45 (Beckman Coulter. c.J33), CD4 (Invitrogen, c.RPA-T4), CD8 (Invitrogen, c.RPA-T8), CD3 (Beckman Coulter, c.UCHTI), CD45RO (Invitrogen, c.HI100), and CCR7 (Miltenyi Biotec, c.REA546). CAR T cell persistence after infusion in peripheral blood was monitored using antibodies CD45 (Beckman Coulter. c.J33), CD4 (Invitrogen, c.RPA-T4), CD8 (Invitrogen, c.RPA-T8), CD3 (Beckman Coulter, c.UCHTI), and human tEGFR cetuximab biosimilar (Invitrogen c.Me183). DAPI (Invitrogen, 62248) was used as a viability dye. For PBMCs characterization the StainExpress Immune Cell Composition Cocktail (Milteryi, USA) were used. All antibodies used for in vitro analysis are described in **Supplementary table 2.**

### End points and study procedures

The primary endpoint was safety. Cytokine release syndrome (CRS) and Immune effector cell-associated neurotoxicity syndrome (ICANS) were graded according to the criteria of American Society for Transplantation and Cellular Therapy consensus grading (DOI: 10.1016/j.bbmt.2018.12.758). Immune Effector Cell-associated Hematotoxicity (ICAHT) was graded according to EHA/EBMT consensus (https://doi.org/10.1182/blood.2023020578) and Immune Effector Cell-Associated Hemophagocytic Lymphohistiocytosis-Like Syndrome (IEC-HS) according to Shah N.N. criteria (DOI: 10.1182/blood.2021011898). All other adverse events were graded according to the National Cancer Institute’s Common Terminology Criteria for Adverse Events, version 5.0.

The secondary endpoint represented overall response rate (ORR), complete response (CR), MRD negativity by the means of flow cytometry, progression free survival (PFS) and overall survival (OS). The clinical efficacy of CD19 CAR T-cells was assessed using the 2018 International Workshop on Chronic Lymphocytic Leukemia (iwCLL2018) criteria (53). Minimal or measurable residual disease (MRD) was assessed by MRD-flow cytometry with sensitivity 10-4 (54). CAR T-cells monitoring was based on CD19 labeled protein and Cetuximab-PE. Response assessment of SLL was implemented according to the Lugano classification 2014 (55), using FDG positron emission tomography-computed tomography (PET-CT) or computed tomography (CT) at +28 days and then every 3 months.

### Cell lines and general cell culture

The HEK293T lentivirus packaging (Clontech, USA) was cultured in DMEM (Gibco, USA) supplemented with 10% FBS (Gibco, USA), 100 U/mL penicillin (Gibco, USA), 100 μg/mL streptomycin (Gibco, USA), and 2 mM GlutaMAX (Gibco). The Nalm-6, C-II and Jeko-1 cell lines were cultured in RPMI 1640 (Gibco, USA) supplemented with 10% FBS (Gibco, USA), 100 U/mL penicillin (Gibco, USA), 100 μg/mL streptomycin (Gibco, USA), and 2 mM GlutaMAX (Gibco). The lentivirus packaging HEK293T cell line was purchased from Clontech, USA. The Jeko-1, Nalm-6 and C-II cell lines were purchased from ATCC, USA. The Nalm-6 cell lines were modified by lentiviral transduction to induce stable firefly luciferase (Ffluc) or green fluorescent protein (GFP) expression. Jeko-1 and C-II cells was transduced with IncuCyte NucLight Red Lentivirus Reagent (EF-1 Alpha, Puro) (Sartorius, USA).

T cells from healthy donors were activated and cultured in RPMI 1640 (Gibco, USA) supplemented with 12.5 ng/mL IL-7 (Miltenyi, USA) and 12.5 ng/mL IL-15 (Miltenyi, USA). All cytotoxicity assays were performed in RPMI medium with IL-7/15. Isolated patients CLL cells were cultured in RPMI-1640 medium (Gibco, USA) supplemented with 10 % heat-inactivated fetal bovine serum (FBS; HyClone, Cytiva), 2 mM L-glutamine (Gibco, USA), 100 U/mL penicillin (Gibco, USA), and 100 µg/mL streptomycin (Gibco, USA). Cells were seeded at 3 × 10^6 cells per well in 24-well plates and incubated at 37 °C with 5 % CO₂. All cell lines were repeatedly tested for the presence of mycoplasma contamination with a MycoReport Mycoplasma Detection Kit (Evrogen, Russia).

### Isolation of primary human peripheral blood mononuclear cells

Human peripheral blood mononuclear cells (PBMCs) were isolated from the blood of patients and healthy donors by gradient density centrifugation on Ficoll-Paque (GE Healthcare, USA) according to a standard protocol approved by the local ethics committee of the Dmitry Rogachev National Medical Research Center of Pediatric Hematology, Oncology, and Immunology. All participants provided informed consent. T cells were isolated from human PBMCs with an Untouched Human T-cell Isolation Kit (Invitrogen, USA) and activated with Dynabeads Human T Activator CD3/CD28 (Gibco, USA) at a 1:1 ratio for 3 days. B cells were isolated from PBMCs by magnetic-activated cell sorting (MACS) using a human B cell isolation kit (Miltenyi, USA), following the manufacturer’s protocol.

### Transduction of T cells with pseudoviral particles

Lentiviral particles containing CAR19 were produced by polyethylenimine-mediated cotransfection (Sigma, USA) of HEK293T cells with the corresponding lentiviral CAR plasmids and the packaging plasmids pCMV VSV-G, pRSV-REV, and pMDLg-pRRE (a gift from Dr. I. Verma). Supernatants were collected at 48 hours post transfection and purified by 2 rounds of centrifugation (1st - 300×g, 5 min, RT; 2nd - 4500×g, 5 min, RT). On the 2nd day after isolation, 1×106/mL activated T cells were resuspended in TexMacs media and mixed with lentiviral supernatants and 13 μg/mL Polybrene (Sigma, USA). Dasatinib (50 nM, Stemcell, Canada) was added prior to transduction. The cells were centrifuged at 1200×g for 90 min at 32 °C and incubated at 37 °C with 5% CO2 for 18 hours. The culture medium was changed on the day after transduction and every 3-4 days to maintain the cells at a density of 2.0×106 cells/mL.

### CLL cell incubation with Lenalidomide

Lenalidomide (Linadeks) was dissolved in DMSO and added to the B-cell cultures at a final concentration of 0.5 µM. Control cells were treated with an equal volume of DMSO. Cells were incubated for 0, 24, or 48 h. At the indicated time points, cells were collected by centrifugation, washed once with PBS, pelleted and stored at −80 °C until further use.

### RNA extraction and quantitative RT-PCR

Total RNA was extracted from frozen B-cell pellets using the RNeasy Mini Kit (Qiagen, Germany) according to the manufacturer’s instructions. RNA concentration and purity were assessed using a NanoDrop 2000 spectrophotometer (Thermo Fisher Scientific, USA). Complementary DNA (cDNA) was synthesized from equal amounts of total RNA using the MINT Reverse Transcription Kit (Evrogen, Russia).

Quantitative PCR was performed using SYBR Green PCR Master Mix (Evrogen, Russia) on a Locus Intero 6 real-time PCR system (Locus Biotechnology). The following cycling conditions were used: initial denaturation at 95 °C for 3 min, followed by 40 cycles of denaturation at 95 °C for 15 s, annealing at 60 °C for 30 s, and extension at 72 °C for 20 s. Gene expression levels were normalized to GAPDH and calculated using the 2^(−ΔΔCt) method.

### CAR T-cell expansion assay

A total of 1×10^5^ CAR19 T cells were cultured in 4 different conditions: + DMSO (control), +1 mkM Ibrutinib (+ibr), +1 mkM Ibrutinib and 10 mkM Lenalidomide (+ibr+len) and +10 mkM Lenalidomide (+len) and were plated in 24-well plates in RPMI medium supplemented with IL-7/IL-15. Cell number and viability were assessed every 2 days for 2 weeks by flow cytometry on an ACEA NovoCyte 2060 (Agilent, USA). ACEA NovoExpress (Agilent, USA) was used for data analysis. On day 2 and 4 cells were stained with CD62L/CD45RA/CD8/CD4 for CAR-T cell differentiation and with Lag-3/Tigit/Tim-3/Pd-1 for T cell exhaustion.

### Cytotoxicity assays of CAR T cells

#### Tumor cells

A total of Jeko-1 or C-II (target cells) were mixed with 1×10^4^ CAR19 cultured in 4 different conditions: + DMSO (control), +1 mkM Ibrutinib (+ibr), +1 mkM Ibrutinib and 10 mkM Lenalidomide (+ibr+len) and +10 mkM Lenalidomide (+len) in different effector to target ratios (E:T ratios were 5:1, 2:1, 1:1). Wells containing only target cells were used as a negative control for cytotoxicity. Wells containing only effector cells were used as a positive control for viability. The cells were incubated at 37 °C with 5% CO2 for 24 hours and washed twice with PBS (Gibco, USA) (300×g, 5 min, RT). The results were evaluated by flow cytometry on an ACEA NovoCyte 2060 (Agilent, USA). ACEA NovoExpress (Agilent, USA) was used for data analysis.

#### CLL patients’ cells and synapse formation

A total of 3×10^5^ CLL cells (target cells) were treated with 10 mkM of lenalidomide (+len) or DMSO (control) for negative control for 48 hours and mixed with non-treated or CAR-T cells cultured in 4 different conditions: + DMSO (control), +1 mkM Ibrutinib (+ibr), +1 mkM Ibrutinib and 10 mkM Lenalidomide (+ibr+len) and +10 mkM Lenalidomide (+len) in (E:T ratios 1:1,1:10, 1:20, 1:50). Cells were incubated at 37 °C with 5% CO2 for 24 hours. Cell suspension from each well was stained with anti-human CD19 and anti-human EGFR and washed twice with PBS (Gibco, USA) (300×g, 5 min, 4 °C). The results were evaluated by flow cytometry using ACEA NovoCyte 2060 (Agilent, USA). ACEA NovoExpress (Agilent, USA) was used for data analysis.

#### Re-challenge assay

A C-II (target cells) were mixed with 1×10^4^ CAR19 cultured in 4 different conditions: + DMSO (control), +1 mkM Ibrutinib (+ibr), +1 mkM Ibrutinib and 10 mkM Lenalidomide (+ibr+len) and +10 mkM (+len) (effector cells) in E:T ratio 3:1. Wells containing only target cells were used as a negative control for cytotoxicity. The cells were incubated at 37 °C with 5% CO_2_ and every 48 hours CAR-T cells were re-challenge with fresh portion of 30×10^4^ C-II cells. On day 6 cells were stained with CD62L/CD45RA/CD8/CD4 for CAR-T cell differentiation and with CAR/LAG-3/TIGIT/TIM-3/PD-1 for T cell exhaustion and the results were evaluated by flow cytometry using ACEA NovoCyte 2060 (Agilent, USA). ACEA NovoExpress (Agilent, USA) was used for data analysis.

### NanoString analysis (RNA)

Primary human T cells were cultivated under different conditions. After 6 days of transduction 500,000 CAR T cells were sorted for EGFRt. 2 days after sorting 1 × 10^6^ CAR T cells were treated with lenalidomide or ibrutinib for 48 hours and 50.000 cells were lysed with RLT buffer. Code-set probes were hybridized with lysed cells for 19 h at 65 °C, subsequently loaded into nCounter MAX cartridges and ran on the nCounter MAX/FLEX according to NanoString protocols. Then, nCounter gene expression assays (NanoString Technologies) were performed using NanoString XT CAR-T Panel Standard. The resulting data were analyzed using nSolver 4.0 software. Downstream bioinformatic analysis was performed with R (4.2.1). Briefly, the differentially regulated genes were identified using the limma R package (56), with a paired design (donor based). Adjusted P value (Benjamini–Hochberg) <0.05 was considered to be significant. The GAGE (57) R package was used to identify regulated gene sets among the the NanoString panel gene sets. P < 0.05 was considered to be significant.

### Enzyme-linked immunosorbent assay (ELISA)

Concentrations of human interleukin-2 (IL-2) and interferon-γ (IFN-γ) in culture supernatants were measured using commercial ELISA kits (Vector-Best, Russia) according to the manufacturer’s instructions. Cell culture supernatants were collected at the indicated time points, centrifuged to remove cell debris, and stored at −80 °C until analysis. Absorbance was measured at the recommended wavelength using a microplate reader. Cytokine concentrations were calculated based on standard curves generated according to the kit protocol.

### Statistical analysis

The frequency and severity of study-related adverse events that are possibly, likely, or definitely related to study treatment, were evaluated for all infused subjects. CR and MRD negativity rate and the associated exact 95% confidence intervals (CIs) were determined among the evaluable subjects. The best overall response rate was denoted as the proportion of patients achieving CR or PR at any timepoint. OS and PFS were evaluated among infused subjects and treated according to the Kaplan-Meier method. Statistics were calculated using Prism Software (version 10.2.0; Graphpad Software Inc., Boston, MA, USA). Statistical analysis was performed using GraphPad Prism 8.0 (GraphPad, USA). Each figure legend denotes the statistical test used. Mean values are plotted as bar graphs, and error bars indicate the standard deviation (SD) unless otherwise stated. ANOVA multiple-comparison p values were calculated using Tukey’s multiple-comparisons test. T test multiple-comparison p values were calculated using the two-stage step-up (Benjamini, Krieger, and Yekutieli) method. All t tests were 2-sided unless otherwise indicated.

## Data Availability

All data produced in the present study are available upon reasonable request to the authors

## Acknowledgements

D.D., A.M. and V.B. were partially supported by Belarusian Republican Foundation for Fundamental Research (Grant No. X25Y-006). V.M.S. and A.G.G. was partially supported by grant no. RSF 25-74-30002.

## Data availability statement

The original contributions presented in the study are included in the article/**Supplementary material**. Further inquiries can be directed to the corresponding author.

## Ethics statement

Ethics committee of Vitebsk Regional Clinical Cancer Centre gave ethical approval for this work. All experiments on human subjects were approved by the D. Rogachev center local ethics committee (decision on 18.01.2018) and were carried out in accordance with the approved protocols. The participants provided their written informed consent to participate in this study.

## Authorship Contributions

Conceptualization and design: M.K., V.M.S., D.D., A.M., A.G.G., M.A.M, and A.V.S. Experiments and data interpretation: All authors. Manuscript drafting: M.K. and V.M.S. Review & Editing: M.K., V.M.S., D.D., A.M., A.G.G., M.A.M, and A.V.S. Final approval of manuscript: All authors. All the authors provided critical and insightful comments.

## Supplementary figures

**Supplementary Table 1.** Final CD19 CAR T-cell products characteristics.

|  |  |  |  |  |  |  |  | CD4+ CAR T-cells subpopulations |  |  |  | CD8+ CAR T-cells subpopulations |  |  |  |  |
| --- | --- | --- | --- | --- | --- | --- | --- | --- | --- | --- | --- | --- | --- | --- | --- | --- |
| Pt ID | Total Cells | Viability % | CD3+CAR + T-cells % | CD4+ T-cells % | CD8+ T-cells % | CD4+CD8 + T cell % | CD4-CD8 - T cell % | TN+ Tscm | TCM | TEM | TEFF | TN+ Tscm | TCM | TEM | TEFF | CD4/ CD8 |
| 01 | 4,00E+09 | 95 | 6 | 15 | 78,9 | 0,93 | 5,17 | 37 | 49,4 | 9,3 | 3,9 | 63 | 25,3 | 6,7 | 5,4 | 0,91 |
| 02 | 1,42E+08 | 85 | 40 | 85,5 | 8 | 2,5 | 4 | 0,62 | 37,18 | 61,78 | 0,42 | 3,87 | 20 | 68,89 | 7,24 | 9,8 |
| 03 | 8,84E+07 | 93 | 61 | 78,3 | 18,3 | 0,3 | 3,1 | 0 | 10 | 89 | 1 | 0 | 3 | 97 | 0 | 5,66 |
| 04 | 3,30E+08 | 89 | 9 | 36,6 | 53,1 | 3,6 | 6,7 | 2,3 | 12 | 85 | 0,7 | 3 | 11 | 85 | 1 | 7,14 |
| 05 | 2,48E+08 | 76 | 22 | 47,7 | 48,4 | 2,23 | 1,67 | 23,3 | 11,6 | 57,6 | 7,5 | 10,9 | 8 | 77 | 4,1 | 0,96 |
| 06 | 1,86E+08 | 90 | 32 | 42 | 52 | 1,3 | 4,7 | 3,1 | 6,4 | 81,6 | 8,8 | 0,7 | 2,1 | 82,3 | 15 | 0,87 |
| 07 | 1,77E+08 | 100 | 35 | 27 | 65 | 4,39 | 3,61 | 0 | 12 | 88 | 0 | 0 | 9 | 91 | 0 | 3,66 |
| 08 | 4,75E+08 | 90 | 29 | 41,9 | 46,1 | 3 | 9 | 8,5 | 9 | 77 | 5 | 5,5 | 14 | 74 | 7 | 1,16 |
| 09 | 2,58E+08 | 100 | 25 | 69,90 | 28,01 | 1,50 | 0,59 | 9,5 | 19 | 68 | 4 | 5 | 13 | 78 | 3,5 | 2,57 |
| 10 | 7,65E+08 | 100 | 4 | 35 | 62 | 2 | 1 | 3,7 | 4,7 | 90 | 2 | 0 | 2,45 | 96,4 | 0,75 | 0,59 |
| 11 | 4,06E+08 | 95 | 25 | 60,4 | 37,5 | 2,06 | 0 | 0,6 | 24,8 | 73 | 1,4 | 2 | 41,3 | 53,4 | 3,5 | 1,6 |
| 12 | 6,66E+08 | 100 | 13 | 72,3 | 23,9 | 1,05 | 2,65 | 3,5 | 50 | 44 | 2 | 4 | 9,5 | 72 | 15 | 2,87 |

CAR T-cells - Chimeric antigen receptor T-cells; TN - Naive T-cells; Tscm - Stem cell memory T-cells; TEM - Effector memory T-cells;

**Supplementary Table 2.** Antibodies for flow cytometry.

|  |  |  |
| --- | --- | --- |
| anti-human CD3-APC | Biolegend | Cat.# 300312 |
| anti-human CD3-FITC | Biolegend | Cat.# 317306 |
| anti-human CD3-APC | Sony | Cat.# 317318 |
| anti-human CD4-FITC | Biolegend | Cat.# 317416 |
| anti-human CD4-FITC | Sony | Cat.# 2102530 |
| anti-human CD4-APC-Cy7 | Thermo Fisher Scientific | Cat.# A15441 |
| anti-human CD4-PE-Cy7 | Sony | Cat.# 2102560 |
| anti-human CD4-BV510 | Sony | Cat.# 2323170 |
| anti-human CD8a-PE | Biolegend | Cat.# 300908 |
| anti-human CD8a-PE | Sony | Cat.# 2105040 |
| anti-human CD8-BV421 | Sony | Cat.# 2323740 |
| anti-human TIGIT-APC | Biolegend | Cat.# 372706 |
| anti-human TIGIT-APC | Sony | Cat.# 2463530 |
| anti-human TIM3-PE | Biolegend | Cat.# 345006 |
| anti-human TIM3-BV510 | Sony | Cat.# 2325150 |
| anti-human LAG3-FITC | Biolegend | Cat.# 369308 |
| anti-human LAG3-BV421 | Sony | Cat.# 2446570 |
| anti-human LAG3-FITC | Sony | Cat.# 2446540 |
| anti-human PD1-BV650 | Sony | Cat.# 2249750 |
| anti-human EGFR-PE | Sony | Cat.# 2364520 |
| anti-human CD25-BV660 | Biolegend | Cat.# 302634 |
| anti-human CD127-BV780 | Biolegend | Cat.# 351330 |

**Supplementary Fig. 2.**
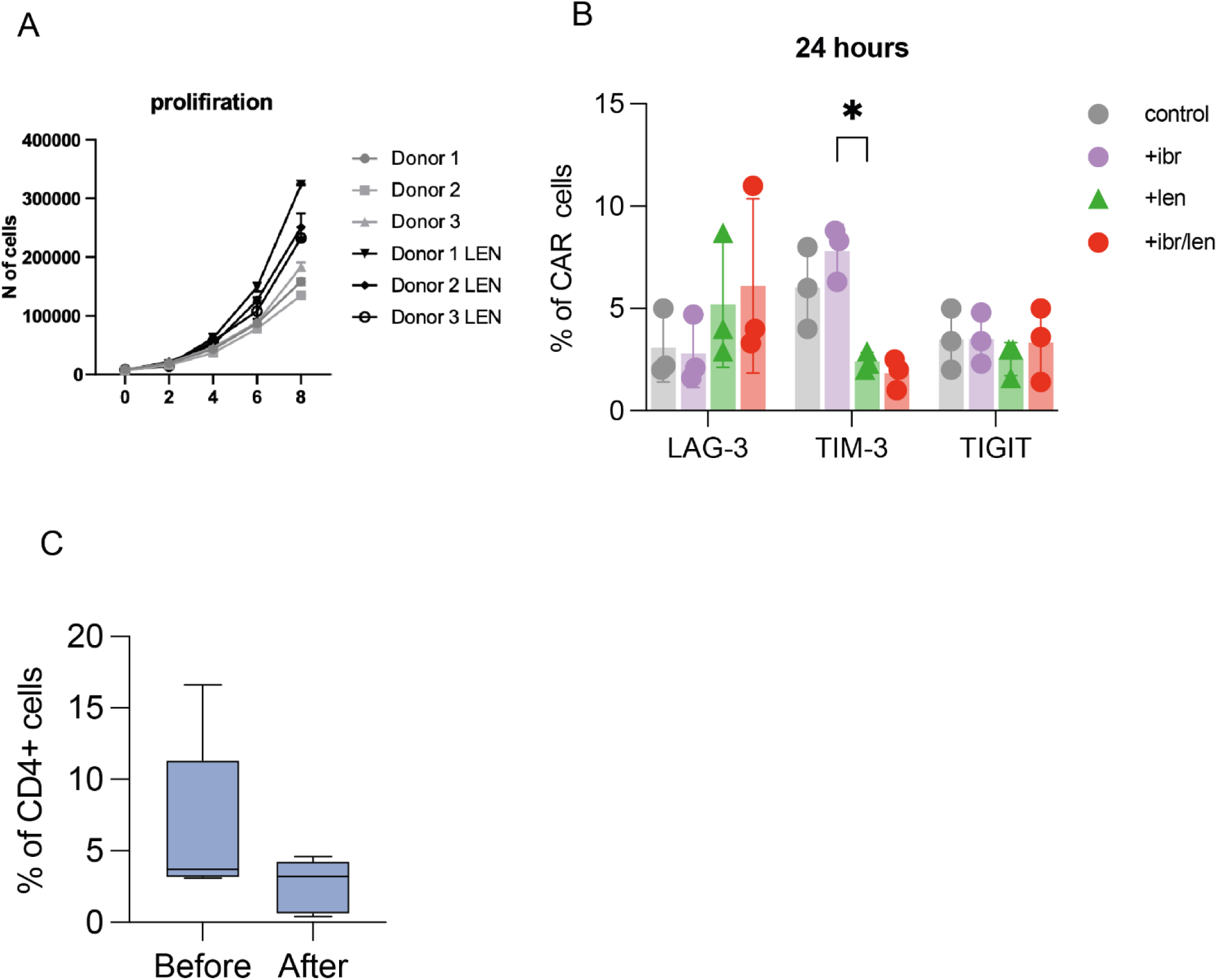
Lenalidomide enhances CAR-T proliferation and cytokine secretion. **A.** Proliferation analysis of CAR-T cells cultured under the indicated treatment conditions for 12 days. **B.** Expression of exhaustion LAG-3, TIM-3, TIGIT markers on day 4, assessed by flow cytometry. Statistical analysis was performed using one-way ANOVA followed by Tukey’s multiple-comparison test. Only significant p values are shown. **C.** Analysis of T reg cells fraction among PBMC isolated from patients before and after lenalidomide cell therapy

**Supplementary Fig. 3.**
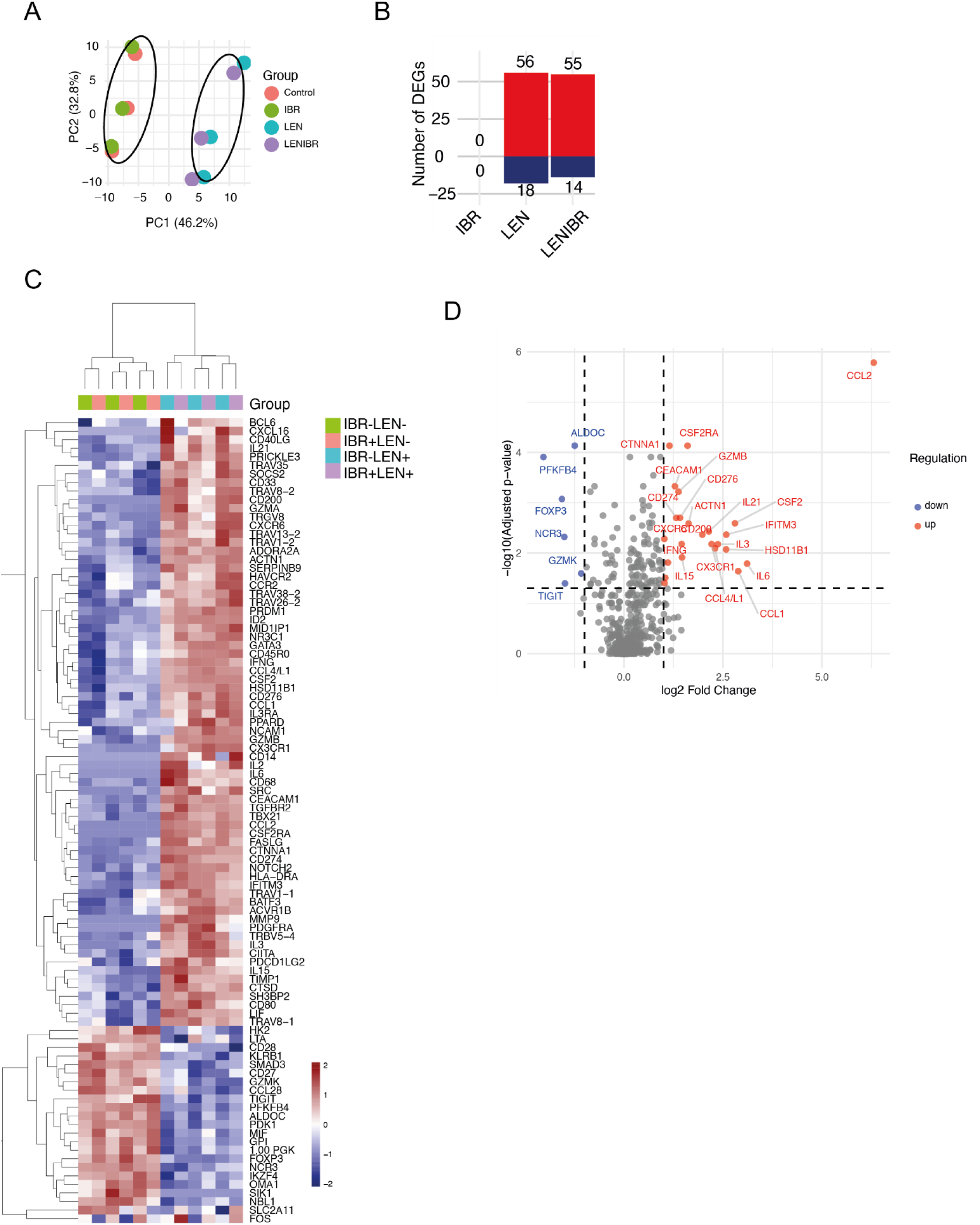
Lenalidomide induces CAR-T transcriptional programs associated with activation and cytotoxicity. **A.** Heatmap of all differentially expressed genes across the indicated treatment conditions. **B.** Volcano plot comparing CAR-T cells treated with ibrutinib alone versus combined ibrutinib and lenalidomide. **C.** Number of differentially expressed genes (DEGs) significantly upregulated and downregulated in each treatment group relative to vehicle control. **D.** Principal component analysis (PCA) of CAR-T cells cultured under the indicated treatment conditions for 24 hours. Three independent donors were analyzed in all panels. Data were generated from CAR-T cells incubated for 24 hours under the indicated conditions. Only significantly differentially expressed genes and pathways are shown, where applicable (P < 0.05).

**Supplementary Figure 4.**
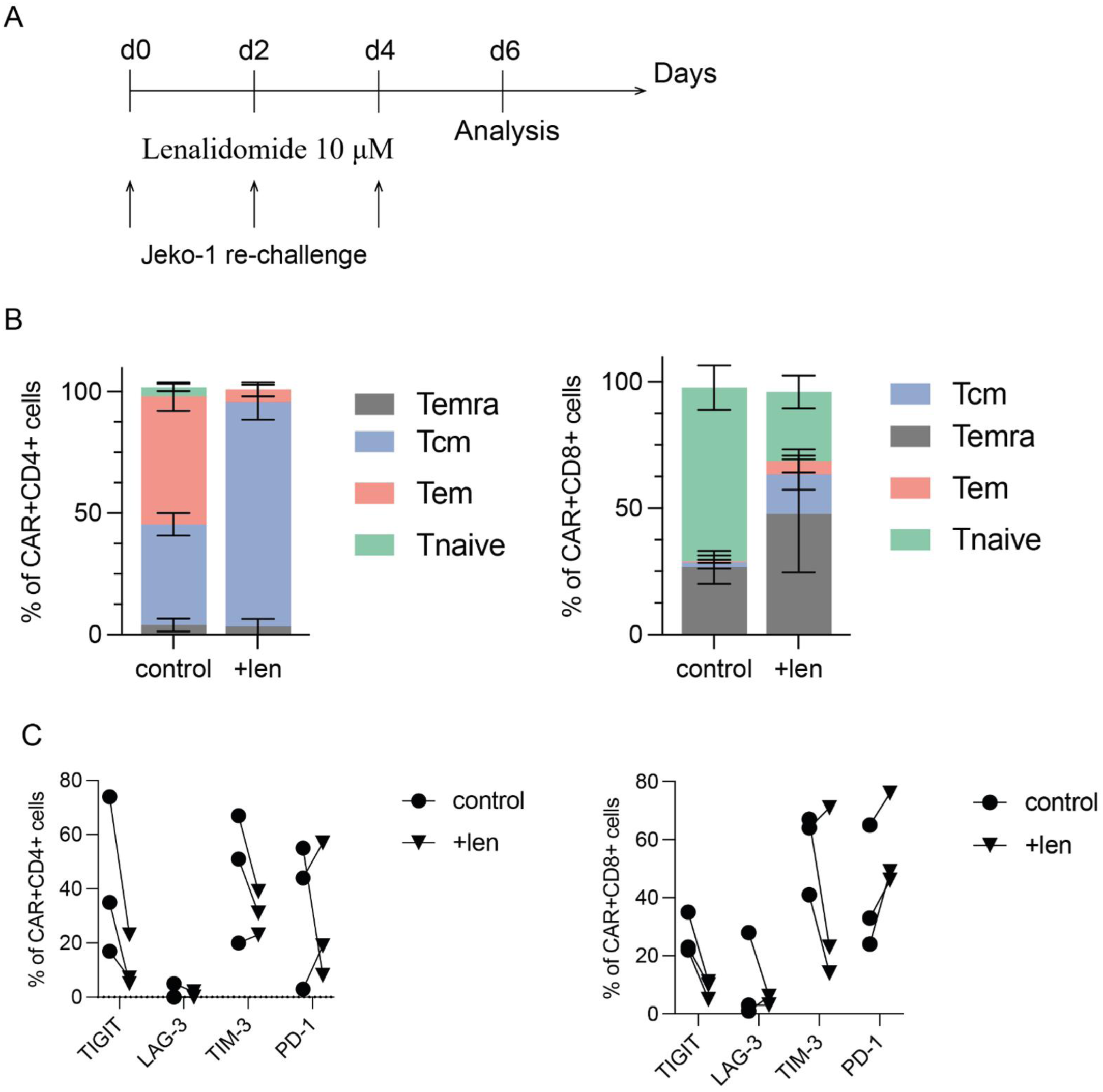
Lenalidomide preserves CAR-T functionality during prolonged antigen challenge. **A.** Experimental design of the prolonged co-culture/rechallenge assay. CD19 CAR-T cells were exposed to lenalidomide or vehicle control and repeatedly challenged with tumor targets to model sustained antigen exposure. **B.** Differentiation phenotype of CAR⁺CD4⁺ and CAR⁺CD8⁺ T cells after prolonged co-culture with tumor targets. T-cell subsets were assessed on day 2 and day 4 by flow cytometry. **C.** Expression of exhaustion-associated markers PD-1, TIM-3, LAG-3, and TIGIT in CAR⁺CD4⁺ and CAR⁺CD8⁺ T-cell subsets on day 4 after prolonged antigen exposure. Lenalidomide-treated CAR-T cells maintained a more favorable phenotype during repeated tumor challenge and showed reduced expression of selected exhaustion-associated markers compared with vehicle-treated controls. Three independent donors were analyzed across panels. Data are shown as mean ± SD. Each dot represents an independent donor. Statistical analysis was performed using repeated-measures two-way ANOVA with Geisser–Greenhouse correction followed by Sidak’s multiple-comparison test. Only significant P values are shown.

**Supplementary Fig. 5.**
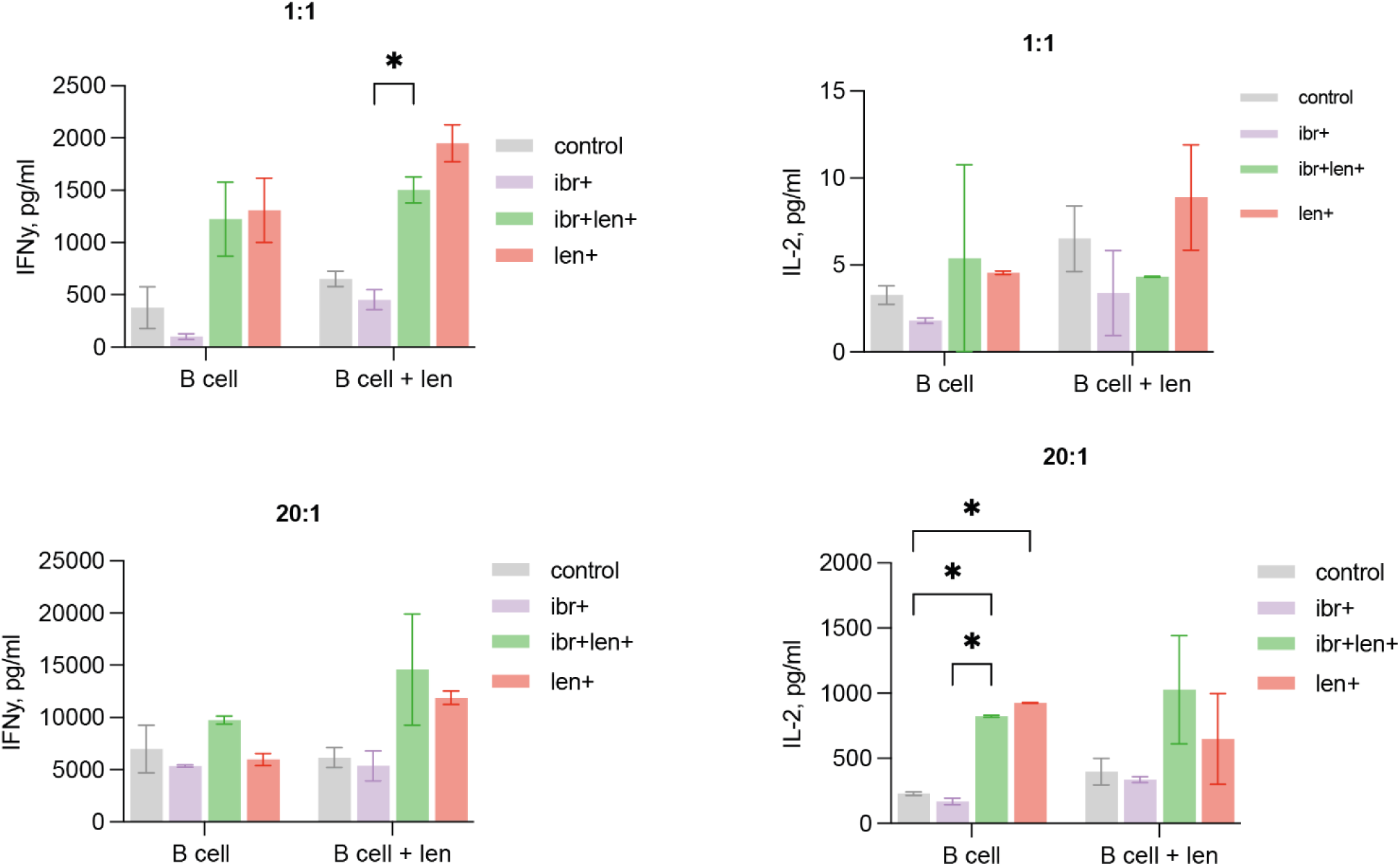
Lenalidomide induces secretion of pro-inflommatory cytokines. IL-2 and IFN-γ secretion by autologous CD19 CAR-T cells after 24-hour co-culture with patient-derived CLL cells. CLL cells were treated with lenalidomide or vehicle control before co-culture. Non-transduced T cells were included as a negative control. Statistical analysis was performed using two-way ANOVA with Geisser–Greenhouse correction followed by Sidak’s multiple-comparison test. Only significant p values are shown.

**Supplementary Fig. 6.**
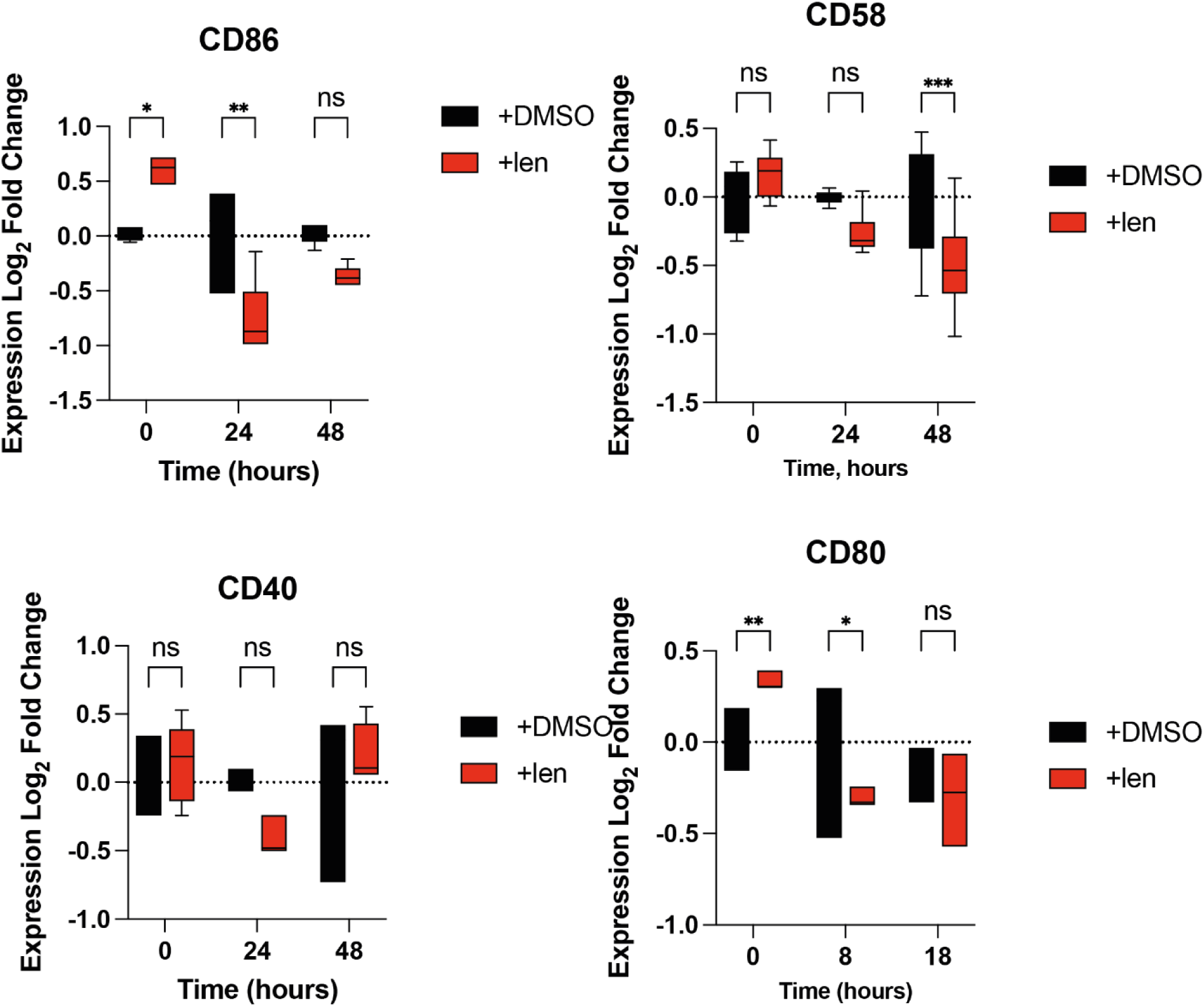
Lenalidomide selectively upregulates CD54/ICAM-1 in patient-derived CLL cells. RT-qPCR analysis of classical costimulatory and adhesion-associated molecules CD86, CD80, CD40, and CD58/LFA-3 in patient-derived CLL cells cultured with lenalidomide or vehicle control for 0, 24, or 48 hours. Lenalidomide induced time-dependent upregulation of CD54/ICAM-1, while CD80, CD86, and CD40 were not broadly induced. CD58/LFA-3 showed variable or reduced expression after lenalidomide exposure. These findings suggest that lenalidomide preferentially remodels the adhesive target-cell interface rather than broadly inducing classical costimulatory molecules. Two independent patient samples were analyzed. Data are shown as mean ± SD. Statistical analysis was performed using repeated-measures two-way ANOVA with Geisser–Greenhouse correction followed by Sidak’s multiple-comparison test. Only significant P values are shown.

## References

1. Hayama M, Riches JC. Taking the Next Step in Double Refractory Disease: Current and Future Treatment Strategies for Chronic Lymphocytic Leukemia. OncoTargets Ther. 2024 Mar 8;17:181–98. doi:10.2147/OTT.S443924

2. Lew TE, Lin VS, Cliff ER, Blombery P, Thompson ER, Handunnetti SM, et al. Outcomes of patients with CLL sequentially resistant to both BCL2 and BTK inhibition. Blood Adv. 2021 Oct 26;5(20):4054–8. doi:10.1182/bloodadvances.2021005083 PubMed PMID: 34478505; PubMed Central PMCID: PMC8945613.

3. Cappell KM, Kochenderfer JN. Long-term outcomes following CAR T cell therapy: what we know so far. Nat Rev Clin Oncol. 2023 Jun;20(6):359–71. doi:10.1038/s41571-023-00754-1 PubMed PMID: 37055515; PubMed Central PMCID: PMC10100620.

4. Jacobs R, Wierda W. Improving Treatment Options for Patients with Double Refractory CLL. Cancers. 2025 Jan 27;17(3):430. doi:10.3390/cancers17030430 PubMed PMID: 39941798; PubMed Central PMCID: PMC11816331.

5. Siddiqi T, Maloney DG, Kenderian SS, Brander DM, Dorritie K, Soumerai J, et al. Lisocabtagene Maraleucel (liso-cel) in R/R CLL/SLL: 24-Month Median Follow-up of TRANSCEND CLL 004. Blood. 2023 Nov 2;142(Supplement 1):330. doi:10.1182/blood-2023-179529

6. Melenhorst JJ, Chen GM, Wang M, Porter DL, Chen C, Collins MA, et al. Decade-long leukaemia remissions with persistence of CD4+ CAR T cells. Nature. 2022 Feb;602(7897):503–9. doi:10.1038/s41586-021-04390-6 PubMed PMID: 35110735; PubMed Central PMCID: PMC9166916.

7. Collins M, Kong W, Jung I, Wang M, Lundh SM, June CH, et al. A Failure to Start: Aborted Activation of CAR T Cells in Chronic Lymphocytic Leukemia. Blood. 2019 Nov 13;134(Supplement_1):681. doi:10.1182/blood-2019-122063

8. Herman SEM, Mustafa RZ, Jones J, Wong DH, Farooqui M, Wiestner A. Treatment with Ibrutinib Inhibits BTK- and VLA-4-Dependent Adhesion of Chronic Lymphocytic Leukemia Cells In Vivo. Clin Cancer Res Off J Am Assoc Cancer Res. 2015 Oct 15;21(20):4642–51. doi:10.1158/1078-0432.CCR-15-0781 PubMed PMID: 26089373; PubMed Central PMCID: PMC4609275.

9. Hartmann TN. Ibrutinib and the chemotactic lymph node choreography. Haematologica. 2024;109(3):698–700. doi:10.3324/haematol.2023.283651

10. Ramsay AG, Clear AJ, Fatah R, Gribben JG. Multiple inhibitory ligands induce impaired T-cell immunologic synapse function in chronic lymphocytic leukemia that can be blocked with lenalidomide: establishing a reversible immune evasion mechanism in human cancer. Blood. 2012 Aug 16;120(7):1412–21. doi:10.1182/blood-2012-02-411678 PubMed PMID: 22547582; PubMed Central PMCID: PMC3423779.

11. Fraietta JA, Lacey SF, Orlando EJ, Pruteanu-Malinici I, Gohil M, Lundh S, et al. Determinants of response and resistance to CD19 chimeric antigen receptor (CAR) T cell therapy of chronic lymphocytic leukemia. Nat Med. 2018 May;24(5):563–71. doi:10.1038/s41591-018-0010-1

12. Krämer I, Engelhardt M, Fichtner S, Neuber B, Medenhoff S, Bertsch U, et al. Lenalidomide enhances myeloma-specific T-cell responses in vivo and in vitro. Oncoimmunology. 2016 Feb 18;5(5):e1139662. doi:10.1080/2162402X.2016.1139662 PubMed PMID: 27467960; PubMed Central PMCID: PMC4910703.

13. Krönke J, Udeshi ND, Narla A, Grauman P, Hurst SN, McConkey M, et al. Lenalidomide Causes Selective Degradation of IKZF1 and IKZF3 in Multiple Myeloma Cells. Science. 2014 Jan 17;343(6168):301–5. doi:10.1126/science.1244851

14. Anderson KC. Lenalidomide and thalidomide: mechanisms of action--similarities and differences. Semin Hematol. 2005 Oct;42(4 Suppl 4):S3–8. doi:10.1053/j.seminhematol.2005.10.001 PubMed PMID: 16344099.

15. Minson A, Hamad N, Cheah CY, Tam C, Blombery P, Westerman D, et al. CAR T cells and time-limited ibrutinib as treatment for relapsed/refractory mantle cell lymphoma: the phase 2 TARMAC study. Blood. 2024 Feb 22;143(8):673–84. doi:10.1182/blood.2023021306

16. Fraietta JA, Beckwith KA, Patel PR, Ruella M, Zheng Z, Barrett DM, et al. Ibrutinib enhances chimeric antigen receptor T-cell engraftment and efficacy in leukemia. Blood. 2016 Mar 3;127(9):1117–27. doi:10.1182/blood-2015-11-679134 PubMed PMID: 26813675; PubMed Central PMCID: PMC4778162.

17. Wang X, Walter M, Urak R, Weng L, Huynh C, Lim L, et al. Lenalidomide enhances the function of CS1 chimeric antigen receptor redirected T cells against multiple myeloma. Clin Cancer Res Off J Am Assoc Cancer Res. 2018 Jan 1;24(1):106–19. doi:10.1158/1078-0432.CCR-17-0344 PubMed PMID: 29061640; PubMed Central PMCID: PMC5991104.

18. Siddiqi T, Soumerai JD, Dorritie KA, Stephens DM, Riedell PA, Arnason J, et al. Phase 1 TRANSCEND CLL 004 study of lisocabtagene maraleucel in patients with relapsed/refractory CLL or SLL. Blood. 2022 Mar 24;139(12):1794–806. doi:10.1182/blood.2021011895

19. Jacobs R, Wierda W. Improving Treatment Options for Patients with Double Refractory CLL. Cancers. 2025 Jan;17(3):430. doi:10.3390/cancers17030430

20. van Bruggen JAC, Peters FS, Mes M, Rietveld JM, Cerretani E, Cretenet G, et al. T-cell dysfunction in CLL is mediated through expression of Siglec-10 ligands CD24 and CD52 on CLL cells. Blood Adv. 2024 Sep 10;8(17):4633–46. doi:10.1182/bloodadvances.2023011934 PubMed PMID: 39042920; PubMed Central PMCID: PMC11401197.

21. Collins MA, Jung IY, Zhao Z, Apodaca K, Kong W, Lundh S, et al. Enhanced Costimulatory Signaling Improves CAR T-cell Effector Responses in CLL. Cancer Res Commun. 2022 Sep;2(9):1089–103. doi:10.1158/2767-9764.CRC-22-0200 PubMed PMID: 36922932; PubMed Central PMCID: PMC10010331.

22. Hartmann TN. Ibrutinib and the chemotactic lymph node choreography. Haematologica. 2024;109(3):698–700. doi:10.3324/haematol.2023.283651

23. Timofeeva N, Gandhi V. Ibrutinib combinations in CLL therapy: scientific rationale and clinical results. Blood Cancer J. 2021 Apr 29;11(4):79. doi:10.1038/s41408-021-00467-7

24. Gauthier J, Hirayama AV, Hay KA, Li D, Lymp J, Sheih A, et al. Comparison of Efficacy and Toxicity of CD19-Specific Chimeric Antigen Receptor T-Cells Alone or in Combination with Ibrutinib for Relapsed and/or Refractory CLL. Blood. 2018 Nov 29;132(Supplement 1):299. doi:10.1182/blood-2018-99-111061

25. Yin Q, Sivina M, Robins H, Yusko E, Vignali M, O’Brien S, et al. Ibrutinib Therapy Increases T Cell Repertoire Diversity in Patients with Chronic Lymphocytic Leukemia. J Immunol. 2017 Feb 15;198(4):1740–7. doi:10.4049/jimmunol.1601190 PubMed PMID: 28077600; PubMed Central PMCID: PMC5296363.

26. Liu Y, Song Y, Yin Q. Effects of ibrutinib on T-cell immunity in patients with chronic lymphocytic leukemia. Front Immunol. 2022 Aug 19;13. doi:10.3389/fimmu.2022.962552

27. Miklos DB, Riedell PA, Bokun A, Chavez JC, Schuster SJ. Leveraging the Immunomodulatory Potential of Ibrutinib for Improved Outcomes of T Cell-Mediated Therapies of B Cell Malignancies: A Narrative Review. Target Oncol. 2025 Mar 1;20(2):217–34. doi:10.1007/s11523-025-01133-9

28. Long M, Beckwith K, Do P, Mundy BL, Gordon A, Lehman AM, et al. Ibrutinib treatment improves T cell number and function in CLL patients. J Clin Invest. 2017 Aug 1;127(8):3052–64. doi:10.1172/JCI89756 PubMed PMID: 28714866; PubMed Central PMCID: PMC5531425.

29. Fraietta JA, Beckwith KA, Patel PR, Ruella M, Zheng Z, Barrett DM, et al. Ibrutinib enhances chimeric antigen receptor T-cell engraftment and efficacy in leukemia. Blood. 2016 Mar 3;127(9):1117–27. doi:10.1182/blood-2015-11-679134 PubMed PMID: 26813675; PubMed Central PMCID: PMC4778162.

30. Gill S, Vides V, Frey NV, Hexner EO, Metzger S, O’Brien M, et al. Anti-CD19 CAR T cells in combination with ibrutinib for the treatment of chronic lymphocytic leukemia. Blood Adv. 2022 Nov 8;6(21):5774–85. doi:10.1182/bloodadvances.2022007317 PubMed PMID: 35349631; PubMed Central PMCID: PMC9647791.

31. Turtle CJ, Hay KA, Hanafi LA, Li D, Cherian S, Chen X, et al. Durable Molecular Remissions in Chronic Lymphocytic Leukemia Treated With CD19-Specific Chimeric Antigen Receptor-Modified T Cells After Failure of Ibrutinib. J Clin Oncol Off J Am Soc Clin Oncol. 2017 Sep 10;35(26):3010–20. doi:10.1200/JCO.2017.72.8519 PubMed PMID: 28715249; PubMed Central PMCID: PMC5590803.

32. Kater AP, Melenhorst JJ. CAR-T and ibrutinib vs CLL: sequential or simultaneous? Blood. 2020 May 7;135(19):1611–2. doi:10.1182/blood.2020005362 PubMed PMID: 32379875.

33. Hanna BS, Yazdanparast H, Demerdash Y, Roessner PM, Schulz R, Lichter P, et al. Combining ibrutinib and checkpoint blockade improves CD8+ T-cell function and control of chronic lymphocytic leukemia in Em-TCL1 mice. Haematologica. 2021 Apr 1;106(4):968–77. doi:10.3324/haematol.2019.238154 PubMed PMID: 32139435; PubMed Central PMCID: PMC8017821.

34. Cantwell M, Hua T, Pappas J, Kipps TJ. Acquired CD40-ligand deficiency in chronic lymphocytic leukemia. Nat Med. 1997 Sep;3(9):984–9. doi:10.1038/nm0997-984 PubMed PMID: 9288724.

35. von Bergwelt-Baildon M, Maecker B, Schultze J, Gribben JG. CD40 activation: potential for specific immunotherapy in B-CLL. Ann Oncol Off J Eur Soc Med Oncol. 2004 Jun;15(6):853–7. doi:10.1093/annonc/mdh213 PubMed PMID: 15151939.

36. Litzinger MT, Foon KA, Tsang KY, Schlom J, Palena C. Comparative analysis of MVA-CD40L and MVA-TRICOM vectors for enhancing the immunogenicity of chronic lymphocytic leukemia (CLL) cells. Leuk Res. 2010 Oct;34(10):1351–7. doi:10.1016/j.leukres.2009.12.013 PubMed PMID: 20122733; PubMed Central PMCID: PMC2891581.

37. Buhmann R, Nolte A, Westhaus D, Emmerich B, Hallek M. CD40-Activated B-Cell Chronic Lymphocytic Leukemia Cells for Tumor Immunotherapy: Stimulation of Allogeneic Versus Autologous T Cells Generates Different Types of Effector Cells. Blood. 1999 Mar 15;93(6):1992–2002. doi:10.1182/blood.V93.6.1992.406k23_1992_2002

38. Palena C, Foon KA, Panicali D, Yafal AG, Chinsangaram J, Hodge JW, et al. Potential approach to immunotherapy of chronic lymphocytic leukemia (CLL): enhanced immunogenicity of CLL cells via infection with vectors encoding for multiple costimulatory molecules. Blood. 2005 Nov 15;106(10):3515–23. doi:10.1182/blood-2005-03-1214

39. Biagi E, Yvon E, Dotti G, Amrolia PJ, Takahashi S, Popat U, et al. Bystander transfer of functional human CD40 ligand from gene-modified fibroblasts to B-chronic lymphocytic leukemia cells. Hum Gene Ther. 2003 Apr 10;14(6):545–59. doi:10.1089/104303403764539332 PubMed PMID: 12718765.

40. Van den Hove LE, Van Gool SW, Vandenberghe P, Bakkus M, Thielemans K, Boogaerts MA, et al. CD40 triggering of chronic lymphocytic leukemia B cells results in efficient alloantigen presentation and cytotoxic T lymphocyte induction by up-regulation of CD80 and CD86 costimulatory molecules. Leukemia. 1997 Apr;11(4):572–80. doi:10.1038/sj.leu.2400598 PubMed PMID: 9096698.

41. Kato K, Cantwell MJ, Sharma S, Kipps TJ. Gene transfer of CD40-ligand induces autologous immune recognition of chronic lymphocytic leukemia B cells. J Clin Invest. 1998 Mar 1;101(5):1133–41. doi:10.1172/JCI1472 PubMed PMID: 9486984.

42. Lapalombella R, Andritsos L, Liu Q, May SE, Browning R, Pham LV, et al. Lenalidomide treatment promotes CD154 expression on CLL cells and enhances production of antibodies by normal B cells through a PI3-kinase-dependent pathway. Blood. 2010 Apr 1;115(13):2619–29. doi:10.1182/blood-2009-09-242438 PubMed PMID: 19965642; PubMed Central PMCID: PMC2852364.

43. Zhang L, Jin G, Chen Z, Yu C, Li Y, Li Y, et al. Lenalidomide improves the antitumor activity of CAR-T cells directed toward the intracellular Wilms Tumor 1 antigen. Hematology. 2021 Dec;26(1):818–26. doi:10.1080/16078454.2021.1981534 PubMed PMID: 34674611.

44. Otáhal P, Prùková D, Král V, Fabry M, Voèková P, Lateèková L, et al. Lenalidomide enhances antitumor functions of chimeric antigen receptor modified T cells. Oncoimmunology. 2016 Apr;5(4):e1115940. doi:10.1080/2162402X.2015.1115940 PubMed PMID: 27141398; PubMed Central PMCID: PMC4839314.

45. Tettamanti S, Rotiroti MC, Giordano Attianese GMP, Arcangeli S, Zhang R, Banerjee P, et al. Lenalidomide enhances CD23.CAR T cell therapy in chronic lymphocytic leukemia. Leuk Lymphoma. 2022 Jul;63(7):1566–79. doi:10.1080/10428194.2022.2043299 PubMed PMID: 35259043; PubMed Central PMCID: PMC9828187.

46. Sabrina Bertilaccio MT, Tettamanti S, Giordano Attianese GMP, Galletti G, Arcangeli S, Rodriguez TV, et al. Low-Dose Lenalidomide Improves CAR-Based Immunotherapy In CLL By Reverting T-Cell Defects In Vivo. Blood. 2013 Nov 15;122(21):4171. doi:10.1182/blood.V122.21.4171.4171

47. Bae J, Keskin DB, Cowens K, Lee AH, Dranoff G, Munshi NC, et al. Lenalidomide Polarizes Th1-specific Anti-tumor Immune Response and Expands XBP1 Antigen-Specific Central Memory CD3+CD8+ T cells against Various Solid Tumors. J Leuk. 2015 Jun;3(2):178. doi:10.4172/2329-6917.1000178 PubMed PMID: 27668268; PubMed Central PMCID: PMC5032910.

48. Zhou D, Zhu X, Xiao Y. CAR-T cell combination therapies in hematologic malignancies. Exp Hematol Oncol. 2024 Jul 18;13(1):69. doi:10.1186/s40164-024-00536-0 PubMed PMID: 39026380; PubMed Central PMCID: PMC11264744.

49. Lemoine J, Morin F, Di Blasi R, Vercellino L, Cuffel A, Benlachgar N, et al. Lenalidomide Exposure at Time of CAR T-Cells Expansion Enhances Response of Refractory/Relapsed Aggressive Large B-Cell Lymphomas. Blood. 2021 Nov 5;138(Supplement 1):1433. doi:10.1182/blood-2021-151109

50. Agarwal S, Aznar MA, Rech AJ, Good CR, Kuramitsu S, Da T, et al. Deletion of the inhibitory co-receptor CTLA-4 enhances and invigorates chimeric antigen receptor T cells. Immunity. 2023 Oct 10;56(10):2388–2407.e9. doi:10.1016/j.immuni.2023.09.001 PubMed PMID: 37776850; PubMed Central PMCID: PMC10591801.

51. Wierda WG, Dorritie KA, Munoz J, Stephens DM, Solomon SR, Gillenwater HH, et al. Transcend CLL 004: Phase 1 Cohort of Lisocabtagene Maraleucel (liso-cel) in Combination with Ibrutinib for Patients with Relapsed/Refractory (R/R) Chronic Lymphocytic Leukemia/Small Lymphocytic Lymphoma (CLL/SLL). Blood. 2020 Nov 5;136(Supplement 1):39–40. doi:10.1182/blood-2020-140622

52. Katsin M, Dormeshkin D, Migas A, Karas O, Shman T, Serada Y, et al. CD19 CAR T cell therapy BY19 for pediatric and adult patients with relapsed or refractory B cell neoplasms in Belarus: Phase 1 trial. Mol Ther Oncol. 2025 Dec 18;33(4):201081. doi:10.1016/j.omton.2025.201081 PubMed PMID: 41322200; PubMed Central PMCID: PMC12664381.

53. Hallek M, Cheson BD, Catovsky D, Caligaris-Cappio F, Dighiero G, Döhner H, et al. iwCLL guidelines for diagnosis, indications for treatment, response assessment, and supportive management of CLL. Blood. 2018 Jun 21;131(25):2745–60. doi:10.1182/blood-2017-09-806398

54. Rawstron AC, Fazi C, Agathangelidis A, Villamor N, Letestu R, Nomdedeu J, et al. A complementary role of multiparameter flow cytometry and high-throughput sequencing for minimal residual disease detection in chronic lymphocytic leukemia: an European Research Initiative on CLL study. Leukemia. 2016 Apr;30(4):929–36. doi:10.1038/leu.2015.313

55. Cheson BD, Fisher RI, Barrington SF, Cavalli F, Schwartz LH, Zucca E, et al. Recommendations for initial evaluation, staging, and response assessment of Hodgkin and non-Hodgkin lymphoma: the Lugano classification. J Clin Oncol Off J Am Soc Clin Oncol. 2014 Sep 20;32(27):3059–68. doi:10.1200/JCO.2013.54.8800 PubMed PMID: 25113753; PubMed Central PMCID: PMC4979083.

56. Ritchie ME, Phipson B, Wu D, Hu Y, Law CW, Shi W, et al. limma powers differential expression analyses for RNA-sequencing and microarray studies. Nucleic Acids Res. 2015 Apr 20;43(7):e47. doi:10.1093/nar/gkv007 PubMed PMID: 25605792; PubMed Central PMCID: PMC4402510.

57. Luo W, Friedman MS, Shedden K, Hankenson KD, Woolf PJ. GAGE: generally applicable gene set enrichment for pathway analysis. BMC Bioinformatics. 2009 May 27;10(1):161. doi:10.1186/1471-2105-10-161

